# A *Plasmodium vivax* controlled human infection and transmission model to evaluate interventions across the life cycle

**DOI:** 10.64898/2026.06.16.26355753

**Authors:** Tessa J.M. Geraedts, Jeroen Bok, Wouter Graumans, Geert-Jan van Gemert, Freia-Raphaella Lorenz, Markus Gmeiner, Rianne Stoter, Karina Teelen, Kjerstin Lanke, Kevin Bos, Katharine A. Collins, Andrew D.S. Duncan, Sarah E. Silk, Francesca R. Donnellan, Carolyn M. Nielsen, Angela M. Minassian, Simon J. Draper, Matthew B.B. McCall, Teun Bousema, Benjamin Mordmüller

## Abstract

**Background:** *Plasmodium vivax* is an underappreciated cause of malaria disease burden. No reproducible and standardized full life-cycle controlled human malaria infection (CHMI) model to accelerate development of novel interventions is available.

**Methods:** This transmission-CHMI trial was conducted in Nijmegen, Netherlands. Healthy, malaria-naive adults were sequentially enrolled into three cohorts of four and inoculated with the asexual blood-stage isolate PvW1. Primary endpoint was proportion of oocyst-positive laboratory-reared *Anopheles stephensi* mosquitoes. The sequential design allowed for adaptations between cohorts. At parasitemia >10 parasites/µL or symptom onset, participants received oral gametocyte-sparing treatment (GST): mepacrine (Cohort 1 and 3; 100 mg at 0, 8 16 hours, then once daily for 3 days) or piperaquine (Cohort 3; 480 mg single-dose). Transmission was assessed by direct skin feeding (DSF) and membrane feeding assay (DMFA) with and without enrichment of gametocytes. End-of-study treatment was atovaquone-proguanil (1000/400 mg once daily for 3 days). The trial was registered: NL-OMON57011.

**Findings:** Participants were enrolled between September 17, 2024 and March 25, 2025, all (12/12) developed parasitemia and transmitted PvW1 to mosquitoes. No serious adverse events occurred. Most adverse reactions were related to malaria. Mepacrine and piperaquine reduced asexual parasitemia while preserving gametocytemia and transmission. Peak transmission occurred within 3 days after GST and depended on the parasite developmental cycle, with highest gametocyte-infectivity ∼48 h post ring-stage. In Cohort 3, mosquito infection reached 100% in all transmission assays. Median peak oocyst counts were 24 (IQR: 14–31) for DSF, 17 (12–19) for DMFA, and 150 (116–199) for enriched DMFA. A two-fold increase in pre-GST maximal parasitemia was associated with 20 additional oocysts (95% CI 8·6–32) in enriched DMFA. Sporozoites were viable in primary human hepatocytes.

**Interpretation:** A PvW1 transmission-CHMI is reproducible and safe, enabling *P. vivax* sporozoite production, relapse models and evaluation of transmission-blocking interventions.

**Funding:** The OptiViVax project is supported by European Union Horizon Europe programme and UK Research and Innovation (UKRI); Swiss Government’s State Secretariat for Education, Research, and Innovation (SERI); National Institute for Health and Care Research Oxford Biomedical Research Centre (NIHR-BRC).

**Research in context:** *Evidence before this study:* In its Malaria Vaccine Technology Roadmap, the World Health Organization prioritizes *Plasmodium vivax* research and vaccine development alongside *Plasmodium falciparum*. Controlled human malaria infections (CHMI) play an important role in the development of new interventions as they enable early evaluation of new drugs and vaccines in small groups of participants.

*Added value of this study:* To our knowledge this is the first study to achieve consistent transmission of a *P. vivax* clone to mosquitoes following blood-stage *P. vivax* CHMI. Reproducible high-level transmission was observed both in direct feeding assays and after gametocyte enrichment and parameters were identified that improve transmission success. It is also the first *P. vivax* CHMI trial that systematically assessed the use of piperaquine and mepacrine to attenuate asexual replication while preserving transmission-competent gametocytes.

*Implications of all the available evidence:* This study expands the repertoire of *P. vivax* CHMI models and builds on the success and knowledge of prior *P. vivax* and *P. falciparum* CHMIs. This optimised transmission model enables production of highly infected mosquitoes and sporozoites for downstream use with a genetically-defined clone, which is particularly important for *P. vivax* because it cannot be maintained in continuous culture. It also facilitates evaluation of transmission-blocking interventions.

## Introduction

*Plasmodium vivax* is a significant and underappreciated cause of malaria in terms of incidence and disease burden [1]. Unlike *P. falciparum*, which predominates in sub-Saharan Africa (SSA), most *P. vivax* cases occur in South and Southeast Asia and South America [2]. Nevertheless, *P. vivax* is also relevant in SSA, despite high Duffy-negativity, and is a major contributor to malaria burden in parts of the Horn of Africa [2, 3]. *P. vivax* is transmitted during acute and chronic infection and can relapse by activation of dormant liver-stage parasites (hypnozoites), which account for approximately 80% of the clinical cases [4].

Chronic and recurrent *P. vivax* infections can have a causative or aggravating role in a wide range of morbidities, including malnutrition, anemia, renal, circulatory, and pulmonary disease [5–8]. Whereas *P. falciparum* mortality is typically driven by acute severe disease, *P. vivax* mostly increases mortality through recurrent and chronic infection [9]. Despite its burden and associated economic losses no vaccines against *P. vivax* are in late-stage development, causing the World Health Organization to prioritize *P. vivax* research and vaccine development in its Malaria Vaccine Technology Roadmap [10].

Controlled human malaria infection (CHMI) studies enable early evaluation of vaccines and drugs in small numbers of healthy volunteers. *P. falciparum* CHMI is a well-established platform that is widely used, including to assess human-to-mosquito transmission [11–13]. CHMI with *P. vivax* is conducted in a few settings worldwide, but validation and standardization are more difficult to achieve [14, 15]. In fact, much of our understanding of *P. vivax* infection dynamics still derives from early 20th-century immunotherapy studies using malaria parasites [16], underscoring the need for updated models.

Biological differences preclude direct translation from *P. falciparum* CHMI to P. viv*ax*. First, *P. vivax* hypnozoites can complicate long-term follow-up and treatment in CHMI studies. Second, unlike *P. falciparum*, *P. vivax* asexual blood stages cannot be maintained in continuous *in vitro* culture. This makes standardized production of sporozoites from cultured parasites impossible and complicates the development of a reproducible pre-erythrocytic CHMI model using infected mosquitoes or cryopreserved *P. vivax* sporozoites. Third, differences in gametocyte differentiation and lifespan require a distinct approach to CHMI transmission studies (i.e. for assessing interventions that block human-to-mosquito transmission). These species-specific characteristics necessitate tailored *P. vivax* CHMI protocols.

Despite these challenges, there has been significant progress in the development of *P. vivax* CHMI models. This includes the recent cryopreservation of erythrocytes containing a parasite clone, PvW1, from Thailand [15], which can be used as an inoculum to initiate blood-stage infections, alongside two older strains from India (2014) [17] and the Solomon Islands (2012) [18]. Proof-of-concept of a *P. vivax* transmission-CHMI has also been established [17, 19]. However, increasing gametocytemia in order to achieve high-level human-to-mosquito transmission generally also requires higher asexual parasitemia, compromising tolerability. Sufficiently high mosquito infection rates for large-scale production of sporozoites and adequately powered transmission studies to test transmission-blocking interventions (TBI) have thus not yet been achieved.

We aimed to improve on earlier results and reach high-level transmission by using antimalarials that suppress asexual replication without compromising gametocyte development [11, 20–24] in combination with *ex vivo* gametocyte enrichment prior to mosquito feeding [17, 25].

## Methods

### Study Design

This single-center CHMI trial was conducted at the Radboud University Medical Center (Radboudumc) in Nijmegen, Netherlands, to establish and optimize a *P. vivax* human-to-mosquito transmission model. Participants were enrolled sequentially in three cohorts of four participants. The sequential design allowed for the adaptation of procedures and protocols to derive an optimized protocol for Cohort 3. The study was approved by the Dutch Central Committee on Research Involving Human Subjects (CCMO; NL-005553). The trial was registered with the Overview of Medical research in the Netherlands database (NL-OMON57011). The full study protocol is available as Supplementary Appendix.

### Participants

Healthy, malaria-naive males and non-pregnant, non-lactating females aged 18-45 years who were Duffy blood group-positive were enrolled. In accordance with Dutch national blood transfusion guidelines, female participants younger than 45 years were screened for Rhesus c antigen, because the blood-stage inoculum was Rhesus c–positive. Sex and ethnicity data were collected by self-report; the options for sex were male and female. All participants met the eligibility criteria as defined in the study protocol (Supplementary Appendix p 21-22) and provided written informed consent before inclusion.

### Procedures

All participants were inoculated with the blood-stage inoculum PvW1, a *P. vivax* clone from Thailand, previously used to develop a standardized blood-stage CHMI model [26]. From Day 5 post-inoculation (pi) participants were assessed daily by a study clinician. Symptoms and parasitemia were monitored until completion of the end-of-study treatment (EOST), no later than Day 26 pi. Study clinicians were reachable by phone 24 hours a day. There were two additional follow-up visits on Day 35 pi and Day 49 pi to confirm the absence of parasitemia and resolution of symptoms. Full details on blood-sampling and follow-up schedules are described in the protocol (Supplementary Appendix p 67).

On the day of inoculation, the PvW1 blood-stage inoculum was thawed and prepared under strict aseptic conditions as previously described [15]. The expected parasite number in the inoculum was calculated backwards from the parasitemia of the parasite donor (Supplementary Appendix p 33). After Cohort 1, and in contrast to the original protocol, the centrifugation brake setting was progressively reduced during processing to minimize disruption of the pellet and optimize parasite recovery. The final protocol used a brake setting of 5 (brake time of ∼35 s). The final clinical inoculum volume was 5 mL for all cohorts and administered intravenously within 4 h of thawing. To confirm bacterial sterility, the prepared inoculum was cultured in blood culture bottles (BD BACTEC).

Participants received two antimalarial treatments: the gametocyte-sparing treatment (GST), to attenuate asexual parasite replication, and the EOST, to clear all parasites. GST was administered if participants had any density of parasitemia with either fever (>38·5°C) or malaria symptoms interfering with daily activities, or if parasitemia exceeded 10 parasites/µL by thick blood smear (TBS). In Cohort 1, participants received mepacrine (quinacrine), in Cohort 2 piperaquine. GST of Cohort 3 was chosen based on the results of Cohorts 1 and 2. Mepacrine treatment consisted of a 100 mg tablet, every 8 h on the first day, followed by once daily for three days. Piperaquine treatment consisted of a single 480 mg tablet. Intake was directly observed by the study physician, either in person or by review of a participant-recorded video. EOST consisted of atovaquone-proguanil (1000/400 mg daily for three days) and was administered to all participants no later than Day 24 pi. Earlier EOST was to be given in case of serious adverse events (SAEs), grade 3 adverse events (AEs) related to GST, parasitemia exceeding 500 parasites/µL on two consecutive days, when all transmission measurements were completed prior to Day 24 pi or at the discretion of the study physician.

### Outcomes

The primary endpoint was proportion of infected mosquitoes per feeding assay. A mosquito was considered infected if at least one oocyst was detected. The objective was infection of ≥80% of dissected mosquitoes (≥9/11). Per assay, 11 mosquitoes were dissected, resulting in 90% power to detect a proportion of ≥20%. Oocyst counting was performed using 1% mercurochrome staining, six to eight days after mosquito infection. Blood was fed to *Anopheles stephensi* mosquitoes between days 11 and 25 pi via direct skin feeding (50 mosquitoes per assay; up to four assays per participant), direct membrane feeding (120 mosquitoes per assay), or membrane feeding with prior gametocyte enrichment (50 mosquitoes per assay). Both direct membrane feeding with serum replacement [27] and gametocyte enrichment methods (Percoll density centrifugation and magnetic cell sorting; MACS) were performed as described previously (https://dx.doi.org/10.17504/protocols.io.ewov1rbkklr2/v1). Three enrichment methods were evaluated during protocol optimization: Percoll 65% centrifuged at 760*g* or at 1500*g*, and MACS.

The secondary endpoint was the number and severity of AEs related to CHMI or study interventions between inoculation and completion of EOST. Data on both solicited and unsolicited AEs were collected at clinic visits, starting from day of inoculation until Day 49 pi. Solicited AEs were classified as being related to inoculation, parasitemia, or antimalarial treatments. SAEs were monitored throughout the entire study period.

Parasitemia was monitored by TBS and qPCR. TBS was performed daily in real time, qPCR in batches. TBS were read independently by two readers with a 95% lower limit of detection of <10 parasites/µL; if results differed by ≥2-fold, a third reading was performed and the mean of the two most concordant readings was reported. The full protocol can be found in the Supplementary Appendix (p 11-12). qPCR targeting the *P. vivax* 18S rRNA gene was performed using the Luna Universal Probe One-Step RT-qPCR Kit (New England Biolabs) as previously described [15]. Gametocytemia was assessed by RT-qPCR targeting the Pvs25 gene for mature female gametocytes as previously described [28].

Following dissection of salivary glands, sporozoites were counted using a Bürker-Türk counting chamber using a phase contrast light microscope as described [29].

Sporozoite infectivity was assessed *in vitro*. Pooled mosquito salivary glands were dissected 14 days after the infectious blood meal and homogenized. Sporozoites were added onto freshly isolated primary human hepatocytes at a ratio of 1:1. Following eight days of incubation, cultures were fixed and stained with anti-*P. falciparum* GAPDH 13.3 (The European Malaria Reagent Repository), goat anti-mouse Alexa Fluor 647 (ThermoFisher Scientific) and DAPI nuclear dye (Sigma Aldrich) and assessed by fluorescence microscopy.

### Statistical analysis

This was an exploratory CHMI study designed to establish a reproducible transmission model. The sample size of participants was based on previous PvW1 CHMI data demonstrating 100% infection success (95% CI 90·5–100%) and ∼50% occurrence of moderate malaria symptoms at a parasitemia ≥10 parasites/µL [15, 26]. Cohorts proceeded sequentially following review of safety and transmission data.

If no participant in a cohort developed parasitemia ≥10 parasites/µL or if mosquito infection rates were <10%, sample size re-estimation was prespecified, subject to ethical approval. Initiation of subsequent cohorts was contingent on review of safety data. An independent data and safety monitoring board reviewed safety outcomes before continuation.

All analyses were exploratory. Association between parasitemia, gametocytemia, and transmission outcomes were assessed using linear regression models. Parasitemia was log_2_-transformed to allow interpretation of regression coefficients per two-fold increase in parasite density. To compare gametocytemia and transmission between assays and conditions, the Wilcoxon signed-rank test was used. All other analyses were descriptive. All analyses included participants who received the inoculum and had available outcome data; no imputation was performed for missing data. Statistical analyses were performed using R version 4.4.1. A p value <0·05 (two-sided) was considered statistically significant.

### Role of the funding source

The funders of the study had no role in study design, data collection, data analysis, data interpretation, or writing of the report.

## Results

Between Sept 17, 2024 (first participant consented) and May 6, 2024 (last participant visit) 12 malaria-naive participants were enrolled sequentially in three cohorts of four participants and successfully infected with asexual blood stage parasites contained in the PvW1 inoculum (Figure 1). Cohort-specific objectives to optimize transmission were achieved (Figure 1). The effective inoculum dose increased with each cohort as the preparation protocol was adapted. Per participant the estimated inoculated parasite numbers were 140 (Cohort 1), 150 (Cohort 2), and 540 (Cohort 3). Baseline characteristics of the participants are presented in Table 1.

**Figure 1.**
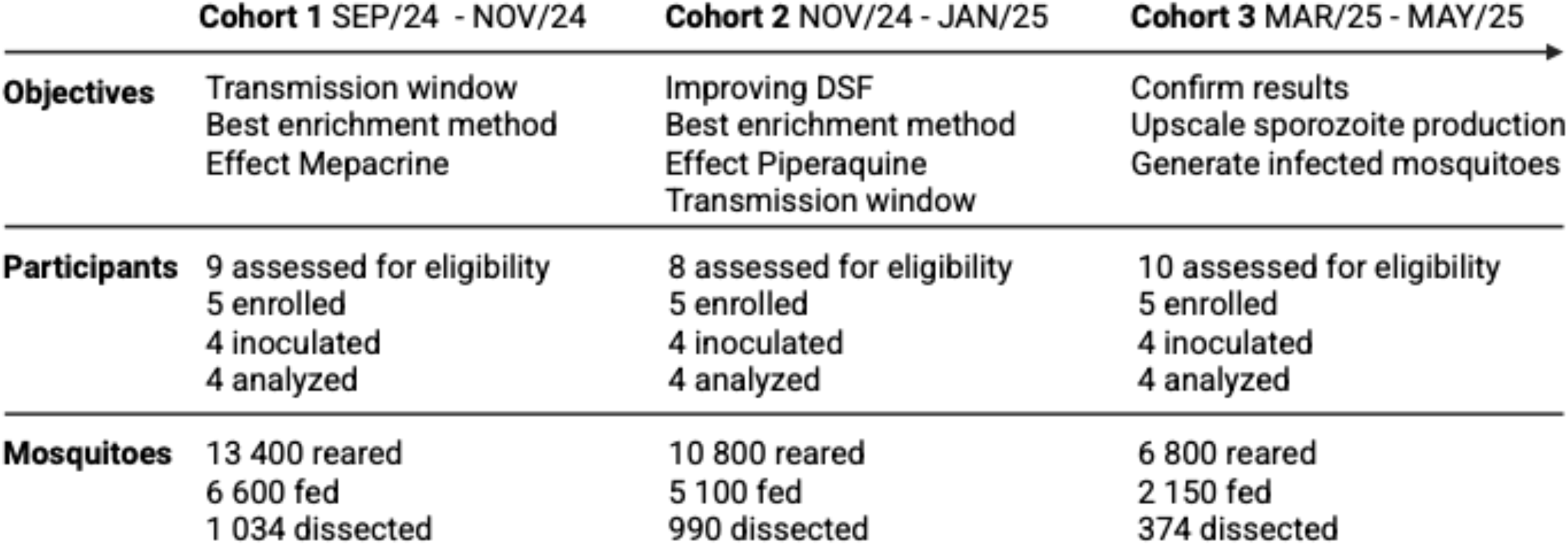
Study Flowchart. Participants were recruited sequentially in three cohorts that differed in their optimization objectives. In total, 27 participants were screened. Reasons for exclusion were: withdrawn consent (n=1), difficult venous access (n=2), negative for Rhesus-c (n=3), conflict of interest (n=1), logistical exclusion (n=2) and conflicting medication (n=3). In total, 13,850 mosquitoes received a blood meal (fed) and 2,398 were dissected to count oocysts or sporozoites. Note that the numbers of processed mosquitoes decreased over time as the window of optimal transmission was identified. DSF= direct skin feeding.

**Figure 2.**
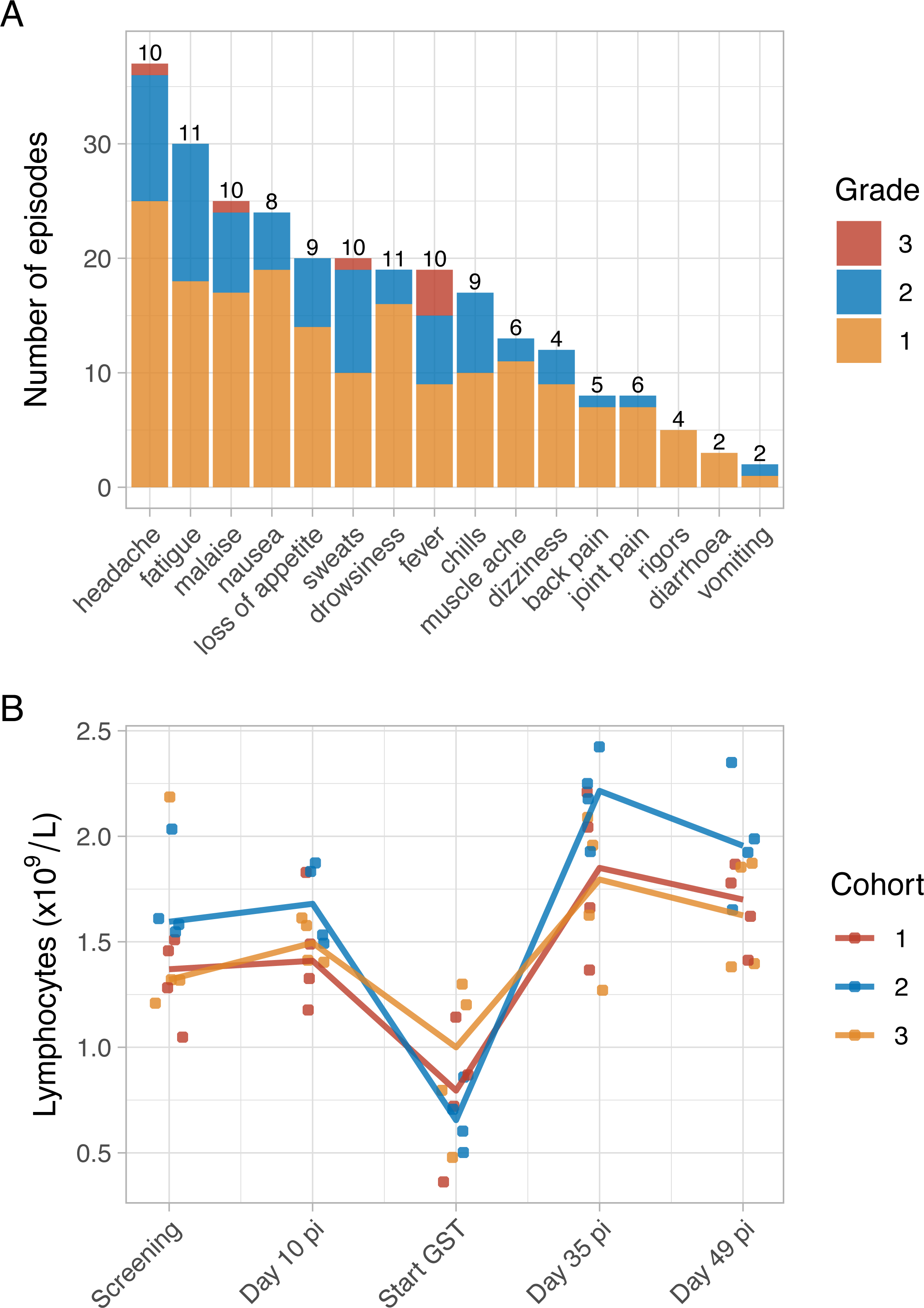
Safety. A) Number of solicited adverse event episodes related to the malaria infection by symptom and severity grade (Grade 1: mild, Grade 2: moderate, Grade 3: severe). Stacked bar chart showing the total number of episodes for each solicited adverse event. Bars are color-coded by grade. The numbers above each bar indicate the number of participants experiencing the symptom at least once out of the total (n = 12). B) Line plot showing the median cohort trajectories of lymphocyte counts over four key study timepoints: Screening Visit, CHMI Day +10, Start GST, CHMI Day +35 and +49. Dots represent individual lymphocyte counts.

**Table 1.**
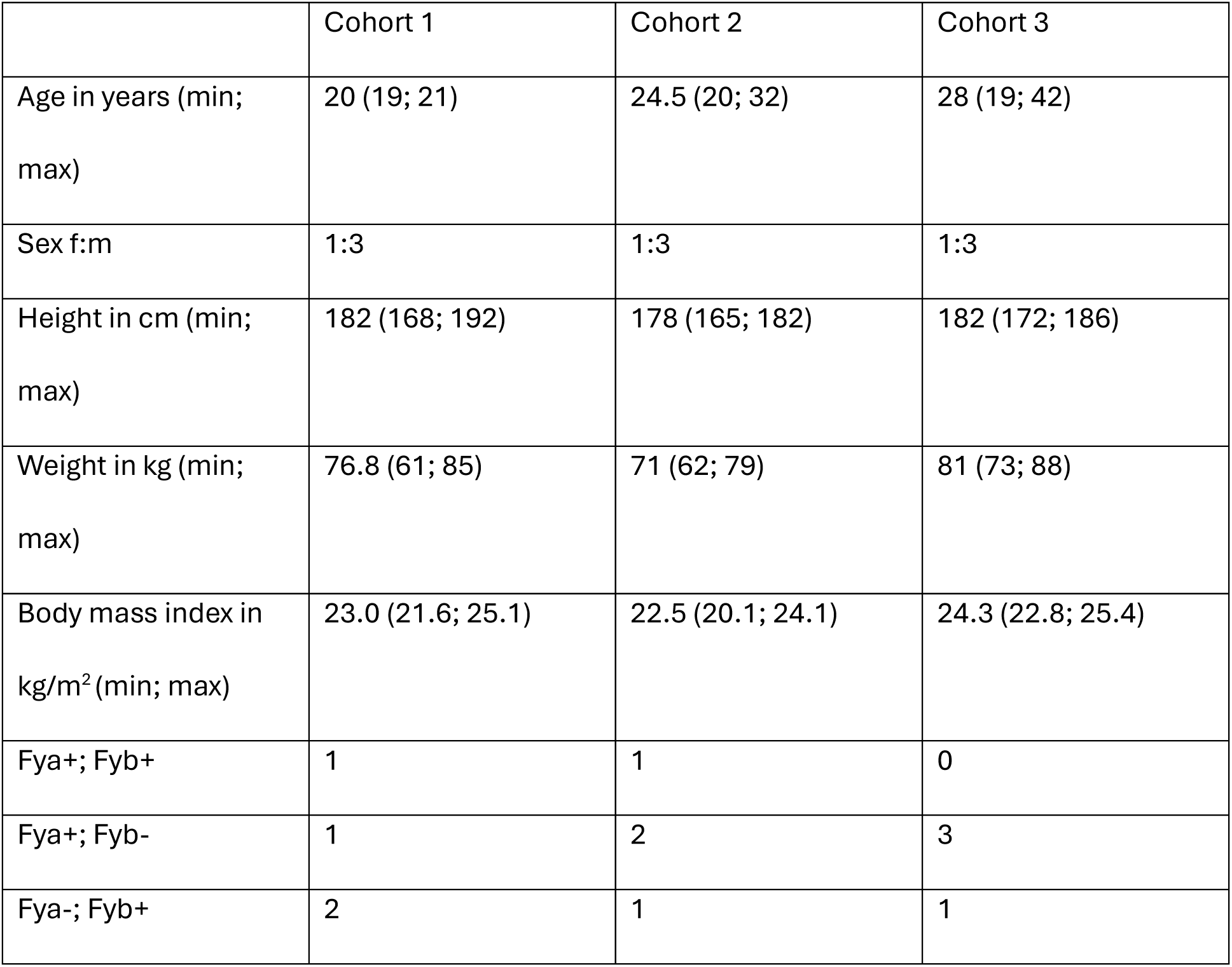
Baseline characteristics of participants. Data are n or mean. Fya and Fyb relates to the expression of the respective Duffy antigens.

|  | Cohort 1 | Cohort 2 | Cohort 3 |
| --- | --- | --- | --- |
| Age in years (min; max) | 20 (19; 21) | 24.5 (20; 32) | 28 (19; 42) |
| Sex f:m | 1:3 | 1:3 | 1:3 |
| Height in cm (min; max) | 182 (168; 192) | 178 (165; 182) | 182 (172; 186) |
| Weight in kg (min; max) | 76.8 (61; 85) | 71 (62; 79) | 81 (73; 88) |
| Body mass index in kg/m<sup>2</sup> (min; max) | 23.0 (21.6; 25.1) | 22.5 (20.1; 24.1) | 24.3 (22.8; 25.4) |
| Fya+; Fyb+ | 1 | 1 | 0 |
| Fya+; Fyb- | 1 | 2 | 3 |
| Fya-; Fyb+ | 2 | 1 | 1 |

GST for Cohort 1 and 3 was mepacrine, for Cohort 2 piperaquine. In Cohort 3 all participants were treated based on the criterion parasitemia ≥10 parasites/μL, whereas all participants in Cohort 2, and 3 out of 4 participants of Cohort 1 were treated because of symptoms affecting daily activities and any parasitemia. One participant of Cohort 1 was parasitemic but did not meet GST treatment criteria. Median times to start GST were similar in Cohort 1 (Day 19 [IQR 18–20]), Cohort 2 (Day 19 [IQR 18–19]) and Cohort 3 (Day 20 [IQR 19–20]). Atovaquone-proguanil (EOST) was initiated between Day 22 pi and Day 24 pi. In Cohort 1, all participants were treated on Day 24 pi. In Cohort 2, two participants were treated on Day 22 pi because of symptom recurrence ≥48 hours after piperaquine initiation. The remaining two participants were treated after all membrane feeding assays (MFAs) had been completed (Day 23). In Cohort 3, all participants were treated on Day 22 pi following completion of all MFAs.

All twelve participants became TBS positive, and parasitemia was cleared in all participants before Day 35 pi. Parasite kinetics are shown in Figure 3. The highest parasitemia measured by TBS was greater in Cohort 3 (484 parasites/µL) compared to Cohort 1 (27 parasites/µL) and Cohort 2 (38 parasites/µL). The median time to first positive 18S qPCR was the same for Cohorts 2 and 3 (Day 13 [IQR 13–13]) but one asexual replication cycle later in Cohort 1 (Day 15 [IQR 14–17]). Parasite multiplication rates (PMR) per cycle were calculated for each cohort using all 18S qPCR measurements above the lower limit of detection (>100 parasites/mL) and prior to treatment. Using this approach, PMR was similar across cohorts: 3·05 (95% CI 0·49–5·61), 4·05 (3·11–4·99), and 4·93 (3·26–6·59) for Cohorts 1–3, respectively. Parasitemia measured by TBS showed a distinct 48-hourly rhythmic pattern with predominantly young rings on uneven days and mature parasites on even days (Figure S1).

**Figure 3.**
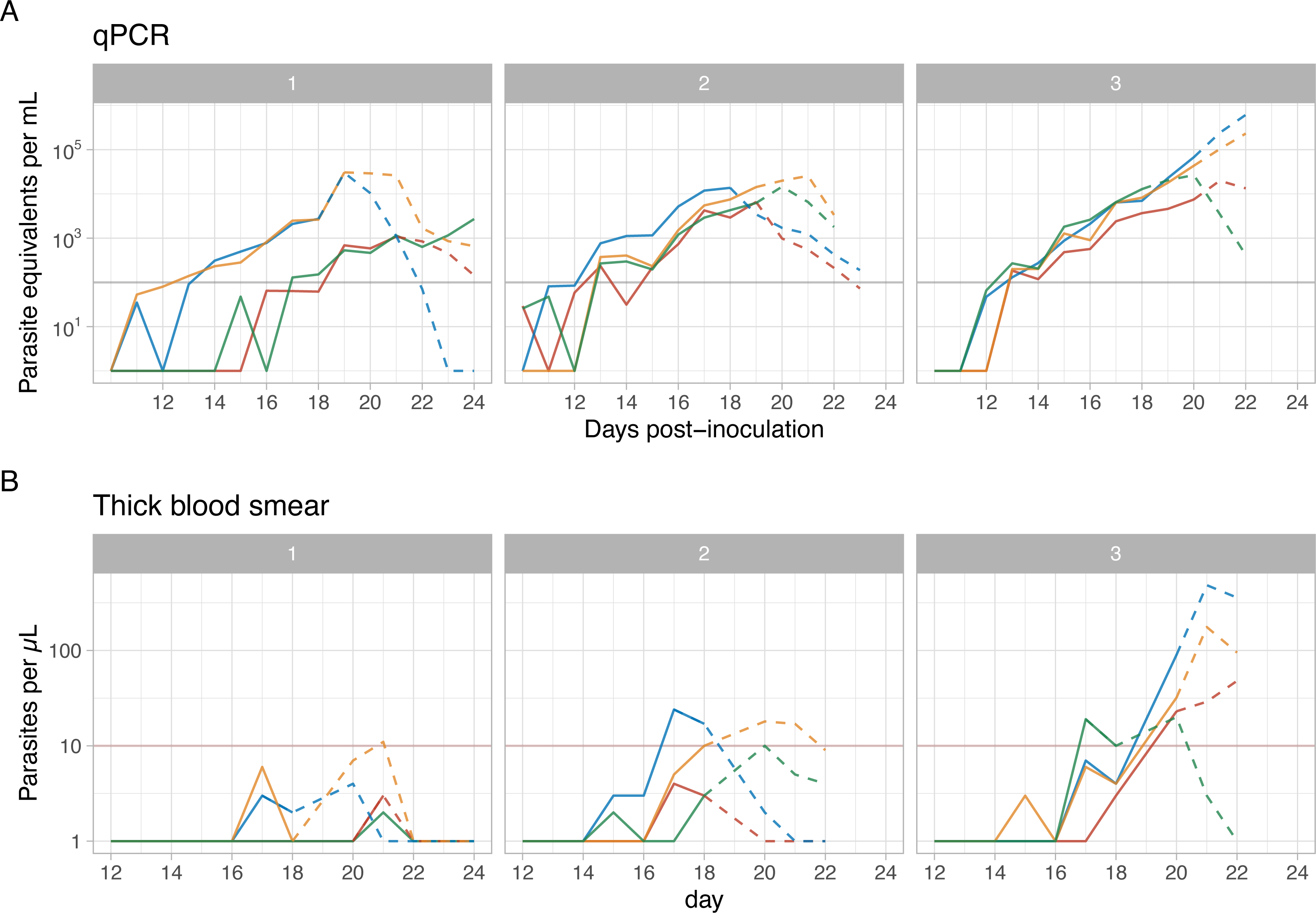
Parasitemia over time. A) 18S qPCR parasitemia (parasites/mL) and B) Thick blood smear counts (parasites/µL) over time in days post-inoculation for each participant. Numbers in grey bars depict the cohorts. The dotted line indicates parasitemia during GST. The horizontal line indicates the threshold for positive result. Measurements stop when end-of-study treatment is initiated.

Gametocytemia was quantified by Pvs25 RT-qPCR and showed a detectable signal in all participants; this continued to increase following start of GST in 10/11 participants (Figure 4a). The participant not requiring GST (Cohort 1) maintained a detectable gametocyte signal 48 hours following start of EOST. To assess the relationship between 18S and Pvs25 RT-qPCR levels, we standardized the values to the mean of 18S and Pvs25 signals in each participant (Figure 4b). As expected, standardized 18S was higher than Pvs25 before GST. 1-3 days following start of GST, this relationship got inverted (Pvs25 > 18S) in all participants receiving GST, indicating that mepacrine and piperaquine acts predominantly on asexual stage parasites. It was noted that the increase in gametocytemia differed between participants starting GST on uneven (rings) and even (trophozoites/schizonts) days (Figure 4c). In the former group gametocytes increased following a lag of one day, while the latter increased immediately (p=0·002, Wilcoxon test).

**Figure 4.**
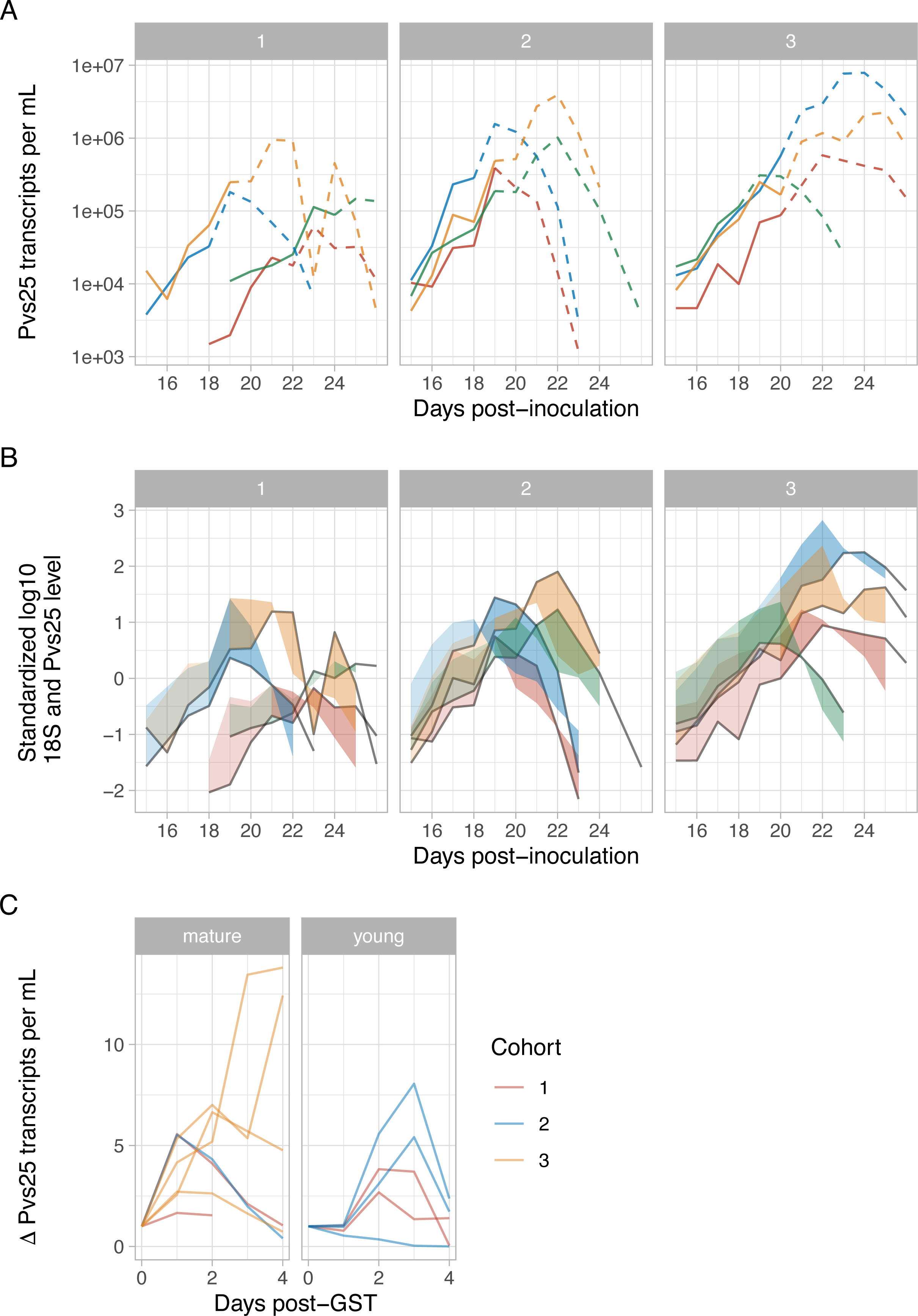
Kinetics of gametocytemia assessed by Pvs25 qPCR. Colors in A) and B) indicate different participants and in C) different cohorts. The three graphs show data of A) Pvs25 transcripts over time, starting with values above the lower limit of detection. The dotted line indicates gametocytemia during GST. B) Standardized transcript-count of Pvs25 and 18S rRNA. The grey line is the Pvs25 and the ribbon the difference to the standardized 18S level. Transparency of the ribbon changes at time of start of GST. Note the change of the ribbon from left of the gametocyte line to the right following start of treatment because asexual parasites are killed while gametocytes continue to circulate. C) Change in log_10_ Pvs25 transcript count after initiation of GST (Day 0). Note that when GST is given during ring stage parasitemia, gametocytes increase with a lag of one day.

Transmission was observed from Day 18 pi onwards (Figure 5a). Highest transmission occurred within 3 days of GST initiation in 11/11 participants. Percoll 65% centrifuged at 1500*g* (Percoll 1500) yielded the highest proportion of infected mosquitoes and oocyst intensities in Cohorts 1 and 2 and was adopted as sole methodology for Cohort 3. This adaptation led to a >2-fold decrease in number of mosquitoes that needed to be used and dissected for Cohort 3 (Figure 1).

**Figure 5.**
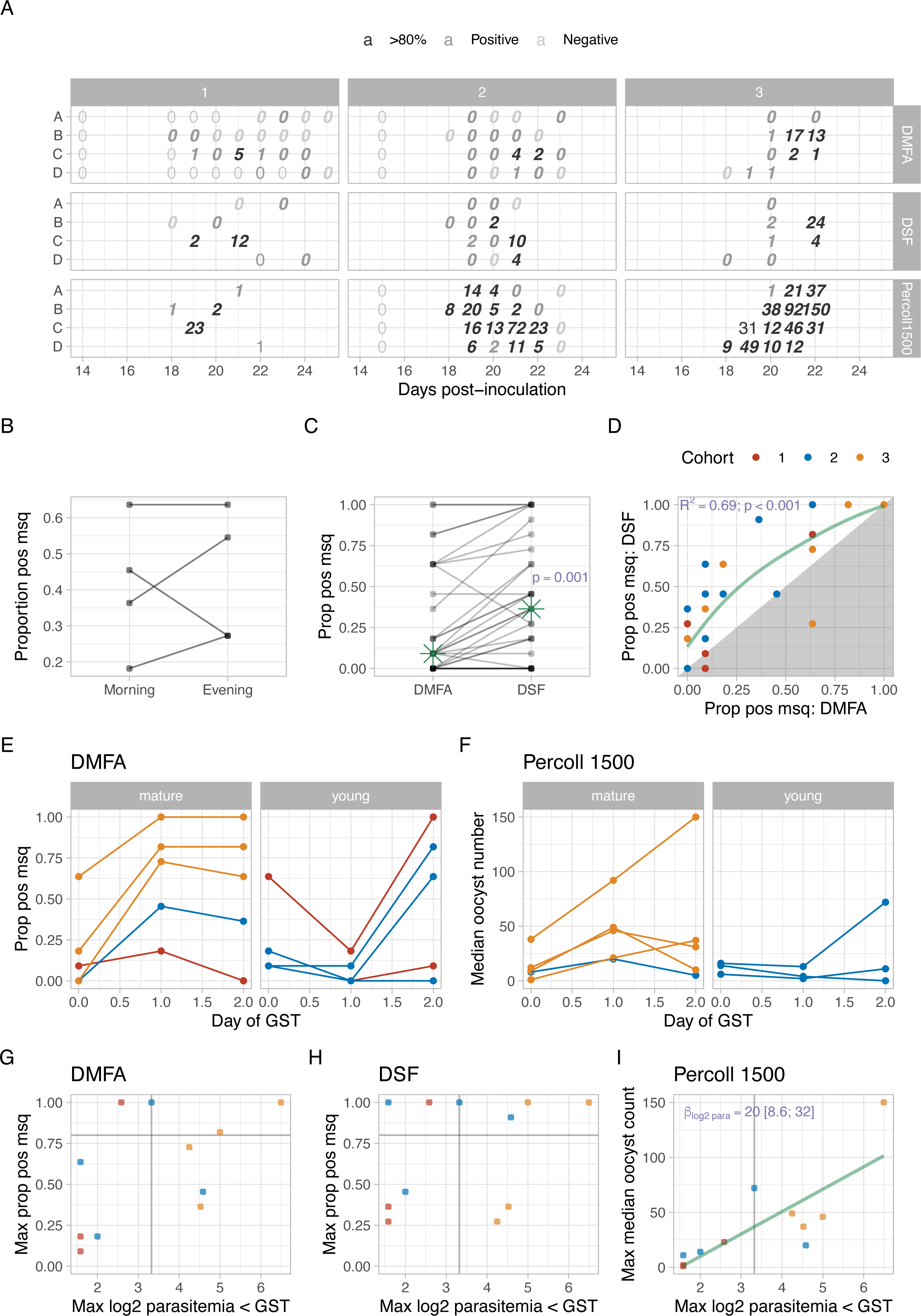
Transmission dynamics of PvW1 over time by different mosquito feeding assays. A) Proportion of infected mosquitoes and median oocyst counts following feeding on blood from individual participants at different timepoints post-inoculation. Columns represent Cohorts (1–3), and assay type (DMFA, DSF, Percoll 1500 enrichment) is shown by panel row. Within each panel, individual participants are arranged along the y-axis. Shading indicates the proportion of infected mosquitoes (light grey = negative; medium grey = positive; dark grey ≥80%). Numbers indicate median oocyst counts. Italicised values indicate measurements obtained during GST. B) Paired comparison of the proportion of infected mosquitoes between morning and evening DSF samples. Each point represents a same-day paired measurement from an individual participant of Cohort 2; lines connect paired samples. C) Paired comparison of the proportion of infected mosquitoes between DMFA and DSF. Each point represents a same-day paired measurement from an individual participant; lines connect paired samples. The green star indicates the median proportion per assay. D) Correlation between the proportion of infected mosquitoes measured by DMFA and DSF. The green line represents the linear regression fit (R²=0·69; p<0·001). Points within the shaded area indicate observations where the proportion was higher by DMFA than DSF. Colors denote cohorts. E) Proportion of infected mosquitoes measured by DMFA before and after initiation of GST. Left panel shows participants treated on days with predominantly mature-stage parasitemia; right panel shows those treated on predominantly young-stage parasitemia days. F) Median oocyst counts following Percoll 1500 enrichment after initiation of GST. (G–H) Association between maximum parasitemia (log₂ parasites/µL) measured by TBS prior to GST initiation and maximum proportion of infected mosquitoes measured by (G) DMFA and (H) DSF. The horizontal grey line indicates 80% mosquito infection; the vertical line indicates 10 parasites/µL by TBS. I) Association between maximum parasitemia prior to GST initiation and median oocyst counts following Percoll 1500 enrichment. The green line represents the linear regression fit. A two-fold increase in maximum parasitemia (log₂ scale) was associated with an increase of 20 median oocysts (95% CI 8·6–32).

Maximum median oocyst count per cage reached 24 (DSF), 17 (DMFA) and 150 (enriched DMFA) oocysts, respectively. In Cohorts 2 and 3, mosquito infection reached >80% across all assays in each participant at least once. In Cohort 2, DSF was performed both in the morning and evening to explore potential diurnal variation in transmission. Mosquito infection rates were similar (Figure 5b). Transmission was consistently higher with DSF compared to unenriched DMFA (Figure 5c) and the proportion of infected mosquitoes closely correlated between the two assays (Figure 5d).

As gametocyte numbers increased differentially according to the timing of GST within the infection cycle, we investigated if the kinetics of transmission followed a similar pattern. Also here, the proportion of infected mosquitoes peaked the day after GST in those treated on (even) days when parasitemia consisted of primarily mature asexual forms, whereas peak transmission lagged by a further day in those treated on (uneven) days when primarily young rings were present. The difference was significant for the proportion of infected mosquitoes in DMFA (p=0·016, Wilcoxon test), as well as for the oocyst count in enriched DMFA for feeds with >80% infected mosquitoes (p=0·036, Wilcoxon test; Figure 5e and f).

We next assessed predictors of transmission intensity to inform optimal sampling timepoints before GST initiation. Besides sampling on uneven days (Figure 5e and f), maximal parasitemia, assessed by TBS before GST initiation, was found to be predictive of transmission (Figure 5g-i). There was no obvious correlation between maximal TBS counts before start of GST and the peak proportion of positive mosquitoes in DMFA or DSF, but a two-fold increase in parasitemia before GST was associated with 20 more oocysts in Percoll 1500 enriched DMFA (95%CI: 8·6-32).

A subset of blood-fed mosquitoes was dissected to count sporozoites in salivary glands and assess their infectivity *in vitro*. Sporozoites became detectable in salivary glands from Day 11 post infectious blood-feed and peaked between Day 14 and 15 (Figure S2). To calculate the number of sporozoites per oocyst, mosquitoes from 15 cages with 100% transmission were dissected for oocyst and salivary sporozoite counts. For oocyst counts, a subset of 11 mosquitoes was dissected per cage, for sporozoites, numbers ranged between 7 and 115 dissected mosquitoes per cage. Weighted linear regression of median oocyst count on sporozoite number on Day 14 post infectious blood-feed resulted in an estimate of 840 salivary gland sporozoites per oocyst (95% CI: 656–1024).

To assess infectivity *in vitro*, freshly isolated primary human hepatocytes were infected with Day 14 salivary gland sporozoites and cultured for eight days. Development of liver stage parasites into schizonts and hypnozoites was confirmed (Figure 6).

**Figure 6.**
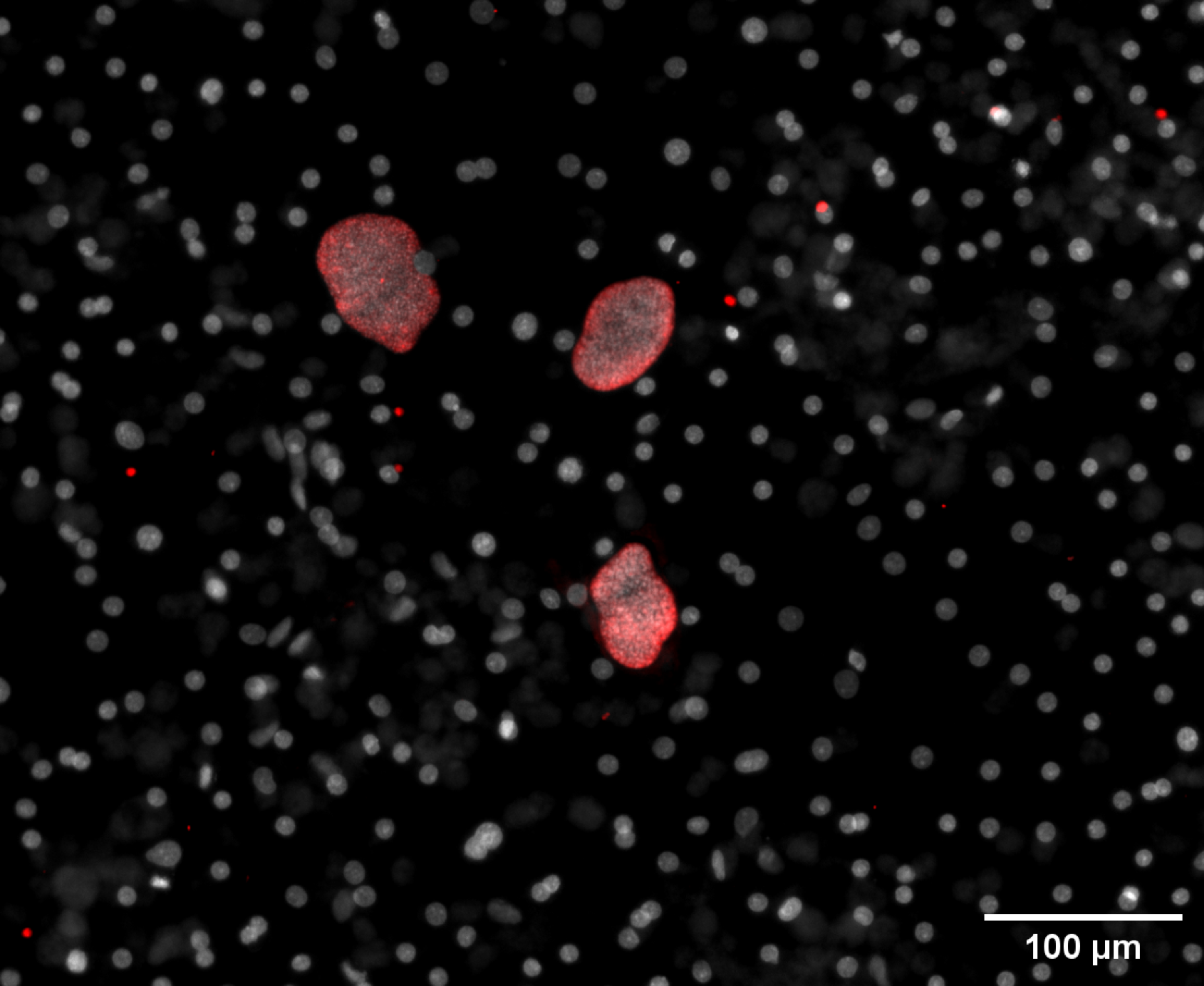
Morphology of liver stage schizonts and hypnozoites in primary human hepatocytes at day 8 post infection. Staining with DAPI nuclear dye (gray) and anti-*Plasmodium* GAPDH (red).

Overall, CHMI was well tolerated; no SAEs occurred, and all participants successfully completed the follow-up. In total, 272 solicited and 85 unsolicited AEs were reported. Every participant experienced at least one AE.

Solicited local AEs related to inoculation were reported in four participants (grade 1, n=5; grade 2, n=1) and included pain, tenderness, erythema, and muscle ache. Four participants reported grade 1 systemic AEs related to inoculation (fatigue or headache). All inoculation-related solicited AEs resolved within 48 hours.

The number and severity of solicited AEs related to malaria are shown in Figure 2a. Five participants reported a grade 3 solicited AE (fever, headache, malaise and sweats). One fever episode lasted for 48 hours, all others resolved within 24 hours. In total, 10/12 participants developed fever during the CHMI. Median time to fever was similar between Cohort 1 (21 [IQR 18–23] days), 2 (19 [IQR 18–19] days) and 3 (19 [IQR 15–20] days). Most solicited AEs attributed to malaria resolved within 48 hours after GST initiation; all symptoms were resolved following EOST.

No AEs were attributed to mepacrine or piperaquine dosing. Two participants in Cohort 2 developed a grade 3 fever two days after piperaquine initiation, indicating low efficacy. This was not observed with mepacrine in Cohort 1. Mepacrine was therefore selected as GST for Cohort 3.

Eight of 12 participants reported solicited AEs related to EOST with atovaquone-proguanil, most occurring on the first day after treatment initiation (Table S1). All AEs were grade 1 (n=24) or grade 2 (n=6). The most frequently reported symptoms were fatigue, headache, and gastrointestinal complaints such as nausea, vomiting, diarrhea, and loss of appetite (Table S1). Of 85 unsolicited AEs 46 were considered related and 39 unrelated to a study procedure. The only severe AE that was related to CHMI procedures was acute allergic urticaria following direct skin feeding (Figure S3). It remained local and resolved within 24 hours with the use of local and systemic antihistamines. Direct skin feeding was otherwise well tolerated, with local skin reactions and itching typical of mosquito bites. Itching was more pronounced and persisted longer in participants in Cohort 2, who underwent four DSFs within three days, compared with two DSFs within three days in Cohort 1 and 3. Three participants had unsolicited AEs (n=4) present at the end of follow-up; all were judged unrelated to CHMI or study interventions. A complete overview of the unsolicited AEs and their MedDRA preferred terms is provided in the Supplementary Appendix (Table S2).

Several laboratory values fluctuated during CHMI. Lymphocyte counts decreased in all participants at the time of GST, with four out of 12 participants having values below the lower reference limit, including two with grade 3 lymphocytopenia (Figure 2b). All counts normalized by Day 35 pi. No participant developed thrombocytopenia. ALT and AST levels tended to increase during CHMI, with grade 1 (ALT n=4; AST n=7) or grade 2 (ALT n=2; AST n=2) elevations (Figure S4). At the last follow-up visit (Day 49 pi), one participant still had a mildly elevated ALT count that was probably related to CHMI and already declining, and another participant had an isolated elevated Gamma-GT, that was considered unlikely related to the CHMI. Both resolved without intervention.

## Discussion

This study established a standardized CHMI transmission model in malaria-naive participants that achieved reproducible high-level transmission of the recently isolated *P. vivax* clone PvW1 to *A. stephensi*. Optimization led to high-levels of transmission using DSF, DMFA and enriched DMFA. Sporozoites harvested from salivary glands infected human hepatocytes *in vitro*, confirming parasite viability and suitability for downstream applications.

Deliberate *P. vivax* infection dates back to early 20th-century malariotherapy, from which much of our current biological understanding derives [16]. Development of the present model was informed by insights from this malariotherapy literature, in addition to pilot enrichment experiments and collaboration with investigators of previous CHMI studies. Recently, modern blood-stage *P. vivax* CHMIs using blood inocula or infectious mosquitoes have been established as platforms for intervention testing [15, 17, 18, 26, 30]. However, development of a standardized full-cycle model including human-to-mosquito transmission has proven more challenging. Two prior studies demonstrated proof of concept. The first achieved transmission to up to 2% of mosquitoes by DMFA and 23% by DSF [19]. The second showed that ≥95% of mosquitoes can be infected through gametocyte enrichment by Percoll density centrifugation while DSF and DMFA results remained comparable to the first study [17, 26]. We reason that consistent (>80% infected mosquitoes) and high-level transmission (>20 oocysts per mosquito) is required to efficiently use the model in clinical development.

In our study, all participants became infectious to mosquitoes. With the optimized protocol (Cohort 3), transmission efficiency exceeded the 80% infected mosquito threshold across all feeding assays, and oocyst intensities surpassed >20 oocysts per mosquito in approximately one-third of MFAs. This high transmission was achieved without compromising tolerability; safety outcomes were comparable to previous CHMIs with PvW1 [15] and other isolates [17,18]. Notably, the different inoculum doses in Cohort 1-3 did not affect the number or severity of AEs.

Beyond potential strain- or site-related differences, the consistent high-level transmission observed was attributable to two adaptations: suppression of asexual parasitemia with mepacrine or piperaquine while preserving transmission-competent gametocytes, and optimization of gametocyte enrichment.

Low-dose piperaquine has been successfully used in *P. falciparum* transmission CHMI [11, 20, 22] and was chosen accordingly. Mepacrine was selected based on malariotherapy data [23, 24]. In our study, both drugs maintained gametocytemia, with transmission increasing after treatment initiation. However, the effect of piperaquine on asexual parasites was transient, consistent with observations in *P. falciparum* CHMI, where recrudescent parasitemia was found after a single dose of 480 mg piperaquine [20]. Mepacrine suppressed asexual parasitemia and symptoms consistently and therefore is currently the best option for GST. At the low-dose regimen used in this study, it was well tolerated. Because all participants received EOST, formal assessment of clinical or parasitological efficacy was not undertaken.

To enrich gametocytes for DMFA we compared MACS and Percoll. In contrast to *P. falciparum* [25], MACS did not increase transmission compared to DMFA. Percoll 65%, particularly at higher centrifugation speed (1500), consistently outperformed DMFA and yielded a high proportion of infected mosquitoes and high oocyst intensities. A more detailed analysis of the different enrichment techniques will be presented elsewhere.

Consistent with known *P. vivax* biology, PvW1 exhibited synchronous 48-hour asexual cycles when examined by TBS, with ring stages on uneven days and mature stages on even days pi. Transmission tended to be higher on uneven days pi and was associated with higher gametocytemia. This pattern suggests that peak transmission depends on the parasite stage at treatment initiation and follows a 48-hour differentiation of rings into gametocytes. Although *P. vivax* gametocyte maturation and lifespan dynamics are not well characterized, malariotherapy data indicate differentiation within 2–3 days and a short gametocyte half-life of ∼1 day [23, 24], consistent with our observations. These findings have practical implications for future studies: if sampling timepoints must be minimized, e.g. for DSF in interventional trials or to perform large scale enriched DMFA, primary mosquito feeding should be focused on the next uneven day following start of GST.

The optimized PvW1 transmission model has two main applications: production of infected mosquitoes and sporozoites for downstream use (e.g., mosquito bite and relapse CHMI and *in vitro* studies), and evaluation of TBIs. High and consistent transmission also facilitates efficient sporozoite production at scale, for example for cryopreservation. For example, to produce 200 vials with 20,000 cryopreserved sporozoites per vial for CHMI, 20 million sporozoites will be needed (assuming an 80% loss due to purification and in-process-tests): 500 mosquitoes with 40,000 sporozoites each, which in turn would require about 100 mL of gametocytemic blood using the current enriched DMFA protocol.

For TBI assessment, the model should approximate natural transmission conditions and incorporate repeated measurements to estimate inhibitory effects. Consistent with previous work, transmission was higher with DSF than unenriched DMFA [17, 31], and both assays correlated. One participant developed a local reaction after their first DSF that precluded further exposure. Strong local reactions have been observed before in about 3% of participants, but systemic anaphylactic reactions have not been reported [32–35]. To maximize output or increase tolerability, DSF can be complemented with DMFA. Given that all participants transmitted and many achieved >80% infection rates, sample size in TBI trials will primarily depend on the minimal detectable effect. As CHMI studies are designed for early “fail-fast” efficacy assessment [13], maximizing statistical power with small cohorts is essential. Limitations reflect the controlled study design: a single *P. vivax* clone was tested in malaria-naive participants, and transmission was assessed using a laboratory-reared mosquito strain. Generalizability to other parasite strains, mosquito strains/sub-species, and *P. vivax*-exposed populations remains to be established, particularly given known variability driven by host- and vector-related factors [36, 37]. E.g., experimental infections conducted in the Brazilian Amazon demonstrated approximately 5-fold species-level differences in infection rates between species when mosquitoes were fed on the same blood meal [38]. Inherent *P. vivax* strain differences can also influence transmission. No direct comparison of strains in malaria-naive participants has been done so far, but it seems that gametocytemia occurs earlier with the isolate HPMV013-Pv than with PvW1 [15, 17]. In addition, we did not include a GST placebo arm, as treatment was administered to enhance safety and tolerability. Finally, gametocyte quantification was based on pvs25 transcripts rather than parasite equivalents, precluding assessment of asexual-to-sexual parasite ratios and gametocyte sex ratios, which may be relevant for TBI studies.

In conclusion, this study establishes a reproducible *P. vivax* CHMI transmission model that balances tolerability and safety with consistent and high transmission. By systematically studying parasite kinetics, treatment- and MFA-timing, and enrichment techniques, we were able to optimize transmission from malaria-naive participants to mosquitoes of a well-defined parasite clone. The model provides a robust framework for evaluating TBIs and enables production of infected mosquitoes and sporozoites for downstream applications, including sporozoite CHMI and pre-erythrocytic vaccine and drug development.

## Supporting information

Full protocol

## Data Availability

All underlying individual, pseudonymized participant data is available on the Radboud Data Repository: https://doi.org/10.34973/01yj-3g97.
Data access will be granted once users have consented to the data sharing agreement and provided written plans and justification for what is proposed with the data. Data access may be obtained by submitting a request in the Radboud Data Repository and will be handled by members of the study team.

## Contributors

TJMG and JB acted as clinical investigators. GvG is the head of the Radboud University Medical Centre Malaria Unit and was responsible for mosquito rearing and mosquito feeding assays. WG oversaw and executed the gametocyte enrichment procedures. RS oversaw TBS preparation and contributed to sample processing. TJMG, JB, FRL, MG, BM, MBBM were certified TBS readers. KT and KL were involved in sample processing and responsible for qPCR data. KB was responsible for processing the PvW1 inoculum. FRL was responsible for the infection of human hepatocytes. MBBM and BM acted as clinical supervisors. BM, TB, AMM, SJD were responsible for the conceptualization of the study. TJMG and BM did the data validation and formal analysis. BM, TB, AMM, SJD acquired funding. MG was the project coordinator. AMM and SJD provided the blood-inoculum PvW1. SES, FRD, CMN shared knowledge and protocols regarding qPCR and processing of the inoculum. KAC shared knowledge and protocols regarding Percoll density gradient centrifugation. TJGM and BM wrote the original draft and final version of the paper. MBBM, TB, ADSD, AMM, SJD, SES, MG, KL, FRL, WG were involved in reviewing and editing. All data was accessible to the authors and they have had final responsibility for the decision to submit for publication.

## Declaration of interests

Several authors received salary support through the funders disclosed in the Acknowledgements. SJD has been a consultant to GSK on malaria vaccines and received additional funding support from Patronus Biotech Pte. AMM has been a consultant to GSK on malaria vaccines and has an immediate family member who received funding support from Patronus Biotech Pte. No other competing interests are declared.

## Data sharing

All underlying individual, pseudonymized participant data is available on the Radboud Data Repository: https://doi.org/10.34973/01yj-3g97.

Data access will be granted once users have consented to the data sharing agreement and provided written plans and justification for what is proposed with the data. Data access may be obtained by submitting a request in the Radboud Data Repository and will be handled by members of the study team.

## Acknowledgments

This study is part of the OptiViVax project and supported by the European Union’s Horizon Europe programme under grant agreement No. 101080744. The OptiViVax project also received co-funding from UK Research and Innovation (UKRI) under the UK government’s Horizon Europe funding guarantee and Swiss Government’s State Secretariat for Education, Research, and Innovation (SERI). The study was also funded in part by the UK National Institute for Health and Care Research Oxford Biomedical Research Centre (NIHR-BRC; the views expressed are those of the authors and not necessarily those of the National Health Service, the NIHR, or the Department of Health).

We thank the study volunteers who participated in this trial. We authors are grateful to Heiman Wertheim as sponsor representative; to Weronika Machnik, Jolanda Klaassen, Laura Pelser-Posthumus and Astrid Pouwelsen for excellent technical assistance; to the Radboud UMC pharmacy, in particular Nicky Lanen-Methley, for helping with regulations regarding drug importation; to all personnel of the Radboud Clinical Research Unit for supporting the clinical investigators; and Annelie Monnier and Annemieke Jansens from Radboudumc, and Fay Nugent, Jee-Sun Cho, Amy Boyd and Phebe Ekregbesi from University of Oxford, for project management support. We also thank Ruth Payne, Quirijn de Mast and Johannes Mischlinger for serving on the Data and Safety Monitoring Board.

Monoclonal antibody anti-GAPDH 13.3 (source: Dr. Jana McBride) was obtained from The European Malaria Reagent Repository (http://www.malariaresearch.eu).

During manuscript preparation, an AI-based language model (ChatGPT 5.4) was used to assist with word-count reduction, prompt: “Edit this text to reduce word count while preserving all scientific meaning. Use concise Lancet Infectious Diseases style. Remove redundancy, shorten sentences, and narrative phrasing without changing content”. The authors take full responsibility for the content of the manuscript.

## Notes

### Clinical Trial

The trial was registered with the Overview of Medical research in the Netherlands database (NL-OMON57011)

### Author Declarations

The study was approved by the Dutch Central Committee on Research Involving Human Subjects (CCMO; NL-005553).

