## Supplementary material for "A *Plasmodium vivax* controlled human infection and transmission model to evaluate interventions across the life cycle": Full protocol

### RESEARCH PROTOCOL

#### Optimizing parasite transmission to laboratory-reared mosquitoes following controlled human malaria infection with *Plasmodium vivax* asexual blood stage parasites in malaria-naïve healthy volunteers in the Netherlands

Protocol version 4.0

18 Decemberr 2024

Optimizing parasite transmission to laboratory-reared mosquitoes following controlled human malaria infection with *Plasmodium vivax* asexual blood stage parasites in malaria-naïve healthy volunteers in the Netherlands

| | |
|---|---|
| **Protocol ID** | RaViCHMI1 |
| **Study register ID** | NL-OMON57011 |
| **Short title** | Radboud Vivax CHMI transmission study |
| **Protocol version** | 4.0 |
| **Protocol date** | 18 December 2024 |
| **Sponsor** | Legal sponsor representative:<br>Heiman Wertheim, MD, PhD<br>Radboudumc<br>Department of Medical Microbiology<br>Tel: +31 (0)24 3614281<br> |
| **Principal investigator** | Benjamin Mordmüller, MD<br>Radboudumc,<br>Department of Medical Microbiology<br>Tel: +31 6 34444464<br> |
| **Project leader** | Matthew McCall, MD, PhD<br>Radboudumc,<br>Department of Medical Microbiology<br>Tel: +31 (0)24 3615363<br> |
| **Subsidising party** | European Union<br>(under Horizon Europe)<br>Grant Agreement: 101080744 |
| **Independent expert** | Arjan van Laarhoven, MD, PhD<br>Radboudumc<br>Department of Internal Medicine<br>Tel: +31 (0)24 361 69 80<br> |
| **Laboratory sites** | Eric Aaldring<br>Radboudumc Clinical Chemical Laboratory<br>Tel: +31 (0)24 3616997<br><br><br>Heiman Wertheim, MD, PhD<br>Clinical Microbiology Laboratory<br>Tel: +31 (0)24 3614281 |

| | |
|---|---|
| | |
| **Pharmacy** | Loek de Jong, PharmD<br>Radboudumc Clinical Pharmacy – Clinical Trial Unit<br>Tel: +31 (0)24-36 17613<br> |
| **Inoculum provider** | University of Oxford<br>Department of Biochemistry<br>South Parks Road<br>OX1 3QU Oxford<br>United Kingdom<br>Tel: +44 (0)1865 613200 |
| **Study protocol contributors** | Jeroen Bok, MD, MSc<br>Teun Bousema, PhD<br>Martin Dresler, PhD<br>Tessa Geraedts, MD<br>Wouter Graumans, PhD<br>Rob ter Heine, PharmD, PhD<br><br>All: Radboudumc |

**DOUCMENT HISTORY**

| Version | Date | Changes |
|---|---|---|
| Version 1 | 24 APR 2024 | - Initial document |
| Version 2 | 02 JUL 2024 | - Document history overview added<br>- Secondary endpoints and number of blood draws aligned throughout the protocol<br>- Additional information on power considerations and gametocyte enrichment procedures added<br>- Data protection approach for data sent outside the European Union added<br>- Editorial changes |
| Version 3 | 30 JUL 2024 | - Argument for use of unauthorized auxiliary medicinal product added |
| Version 4 | 18 DEC 2024 | - Study register ID added<br>- Exclusion criteria added<br>- Appendix 2 updated<br>- Timepoints mosquito feeding assays updated (based on cohort 1)<br>- Clarified the criteria for start end of study treatment before Day+24 |

**PROTOCOL SIGNATURE SHEET**

| Name | Signature | Date |
|---|---|---|
| **Sponsor or legal representative:**<br>**Head of Department:**<br>Heiman Wertheim, MD, PhD<br>Radboudumc<br>Department of Medical Microbiology | | |
| **Principal Investigator:**<br>Benjamin Mordmüller, MD<br>Radboudumc<br>Department of Medical Microbiology | | |

9.2.1. Adverse events (AEs) ... 44
9.2.2. Serious adverse events (SAEs) ... 45
9.2.3. Suspected unexpected serious adverse reactions (SUSARs) ... 45
9.3. Annual safety report ... 46
9.4. Follow-up of adverse events ... 47
9.5. Adverse event data collection ... 47
9.5.1. Recording of adverse event data collection ... 47
9.5.2. Assessment of causality ... 48
9.6. Data Safety Monitoring Board (DSMB) ... 49
10. STATISTICAL ANALYSIS ... 52
10.1. Primary study parameter(s) ... 52
10.2. Secondary study parameter(s) ... 52
10.3. Other study parameters ... 52
10.4. Interim analysis (if applicable) ... 53
11. ETHICAL CONSIDERATIONS ... 54
11.1. Regulation statement ... 54
11.2. Recruitment and consent ... 54
11.3. Objections by minors or incapacitated subjects ... 55
11.4. Benefits and risks assessment, group relatedness ... 55
11.5. Compensation for injury ... 55
11.6. Incentives ... 56
11.7. Other ethical considerations ... 56
11.7.1. Incidental findings ... 56
11.7.2. Environment ... 57
12. ADMINISTRATIVE ASPECTS, MONITORING AND PUBLICATION ... 58
12.1. Handling and storage of data and documents ... 58
12.1.1. Source Data ... 58
12.1.2. Data protection & safeguarding: ... 59
12.2. Monitoring and Quality Assurance ... 61
12.2.1. General considerations ... 61
12.2.2. Study monitoring ... 61
12.2.3. Independent Auditing ... 62
12.2.4. Inspections ... 62
12.3. Amendments ... 63
12.4. Annual progress report ... 63
12.5. Temporary halt and (prematurely) end of study report ... 63
12.6. Public disclosure and publication policy ... 64
13. REFERENCES ... 65
Appendix 1: Schedule of procedures ... 67
Appendix 2: Sampling schedule for measurements depending on initiation of gametocyte-sparing treatment ... 69

#### LIST OF ABBREVIATIONS AND RELEVANT DEFINITIONS

| | |
|---|---|
| **AE** | Adverse Event |
| **β-HCG** | β-Human Chorionic Gonadotropin |
| **CHMI** | Controlled Human Malaria Infection |
| **CRF** | Case Report Form |
| **CYP** | Cytochrome |
| **DFA** | Direct Skin Feeding Assay |
| **DMFA** | Direct Membrane Feeding Assay |
| **DMP** | Data Management Plan |
| **DPO** | Data Protection Officer |
| **DSMB** | Data Safety Monitoring Board |
| **eCRF** | electronic Case Report Form |
| **ECG** | Electrocardiogram |
| **EDC** | Electronic Data Capture |
| **EEG** | Electroencephalogram |
| **EU** | European Union |
| **GCP** | Good Clinical Practice |
| **GDPR** | General Data Protection Regulation; in Dutch: Algemene Verordening Gegevensbescherming (AVG) |
| **GP** | General Practitioner |
| **GST** | Gametocyte Sparing Treatment |
| **HBsAg** | Hepatitis B Surface Antigen |
| **HBV** | Hepatitis B Virus |
| **HCV** | Hepatitis C Virus |
| **HIV** | Human Immunodeficiency Virus |
| **ICD** | International Classification of Disease |
| **ICF** | Informed Consent Form |
| **ICH** | International Council for Harmonisation of Technical Requirements for Pharmaceuticals for Human Use |
| **ICTRP** | International Clinical Trials Registry Platform |
| **IBSM** | Induced Blood Stage Malaria |
| **LLoD** | Lower Limit of Detection |
| **LSM** | Local Safety Monitor |
| **MACS** | Magnetic Activated Cell Sorting |

| | |
|---|---|
| **MedDRA** | Medical Dictionary for Regulatory Activities |
| **METC** | Medical research ethics committee (MREC); in Dutch: medisch-ethische toetsingscommissie (METC) |
| **NFU** | The Netherlands Federation of University Medical Centers; in Dutch: Nederlandse Federatie van Universitair Medische Centra |
| **PBMC** | Peripheral blood mononuclear cells |
| **PCR** | Polymerase Chain Reaction |
| ***P. falciparum*** | *Plasmodium falciparum* |
| **PfSPZ** | *Plasmodium falciparum* Sporozoites |
| **PI** | Principal Investigator |
| ***P. vivax*** | *Plasmodium vivax* |
| **PvW1** | *Plasmodium vivax* W1 |
| **qPCR** | quantitative Polymerase Chain Reaction |
| **Rh** | Rhesus |
| **SAE** | Serious Adverse Event |
| **SOP** | Standard Operating Procedure |
| **Sponsor** | The sponsor is the party that commissions the organisation or performance of the research, for example a pharmaceutical company, academic hospital, scientific organisation or investigator. A party that provides funding for a study but does not commission it is not regarded as the sponsor, but referred to as a subsidising party. |
| **SUSAR** | Suspected Unexpected Serious Adverse Reaction |
| **TBS** | Thick Blood Smear |
| **UPS** | Uninterruptible Power Supply |
| **WHO** | World Health Organization |
| **WMO** | Medical Research Involving Human Subjects Act; in Dutch: Wet Medisch-wetenschappelijk Onderzoek met Mensen |

#### SUMMARY

**Rationale:** *Plasmodium vivax* is the second-most important malaria parasite (following *P. falciparum*) and has a significant disease burden, particularly in South and Southeast Asia as well as in South America. In parts of Africa, *P. vivax* also results in a significant human disease burden. The World Health Organization has therefore included the development of vaccines to prevent infection and transmission of *P. vivax* into its updated Malaria Vaccine Technology Roadmap. Interventions to reduce transmission are particularly important to malaria control as the basic reproduction rate ($R_0$) of *Plasmodium* parasites can exceed 1000. To assess the effect of medicinal interventions on transmission, a controlled human malaria infection (CHMI) model has been established and validated successfully for *P. falciparum*. RaViCHMI1 will adapt the *P. falciparum* CHMI model to measure transmission from humans to mosquitoes for *P. vivax*.

**Objective**:

*Main objective*: Develop a protocol to reproducibly infect mosquitoes with blood from human study participants undergoing asexual blood stage CHMI with the *P. vivax* clone PvW1.

*Secondary objectives*: Maximize mosquito infection rate with PvW1 by using I) *ex vivo* gametocyte concentration techniques, and II) antimalarial treatment that minimizes malaria-associated symptoms and adverse events due to asexual parasitaemia while not affecting gametocyte viability during CHMI (gametocyte-sparing treatment).

**Study design:** Single centre, sequential, non-controlled, non-randomized transmission study.

**Study populations:** I) Healthy, malaria-naïve adult male and female volunteers, 18 to 45 years old, II) Laboratory-reared *Anopheles stephensi* mosquitoes, 1 to 5 days old.

**Intervention**: All participants undergo CHMI with PvW1 and receive gametocyte-sparing and end-of-study antimalarial treatment.

**Main study endpoints:**

*Primary endpoint*: Proportion of infected mosquitoes, measured as oocyst positivity, following exposure to blood of participants with *P. vivax* parasitaemia.

*Secondary endpoint*: Number and severity of adverse events related to CHMI between inoculation and completion of end-of-study treatment.

**Nature and extent of the burden and risks associated with participation, benefit and group relatedness:**

*Inoculum*: Participants will undergo CHMI using a cryopreserved blood inoculum of the *P. vivax* clone PvW1. The inoculum has been produced from a prospectively screened and infected donor with a universal blood group who passes all criteria for blood donation. All procedures related to producing the blood inoculum were done under strict quality assurance. The inoculum has been used safely in 37 malaria-naïve participants in the UK, so far.

*Malaria*: The main risk of *P. vivax* malaria are symptoms of systemic inflammation, such as fatigue, fever, headache, and myalgia. It is commonly characterized as “benign tertian malaria” because, in contrast to *P. falciparum* malaria, the risk of complications in healthy adults with a recently acquired infection is extremely low. Nevertheless, there is a significant disease burden including complications and death, which are associated with chronic and relapsing infections, malaria in pregnancy, and co-morbidities. As a blood inoculum is used, there is no risk of *P. vivax* relapse. The antimalarial treatment is curative, therefore no complications of chronic or recurrent malaria may occur.
*Antimalarial treatment*: Participants will receive gametocyte-sparing antimalarial treatment with piperaquine or mepacrine as well as end-of-study treatment with atovaquone-proguanil or artemether-lumefantrine. All antimalarials have been given in millions of doses and their safety profile is well described. Piperaquine, mepacrine and lumefantrine can prolong the QT interval. Therefore, an electrocardiogram is part of screening. Participants with significant ECG abnormalities will be excluded from participation at the screening visit.
*Mosquito bites*: Mosquito bites can cause local discomfort. Mild local inflammation and pruritus typically accompanies the bite of the insect. In contrast to other insect bites (e.g. bees) anaphylaxis after mosquito bites is extremely rare and has never been reported in CHMI studies.
*Phlebotomy*: There will be 27 scheduled visits with phlebotomies: at screening, before inoculation, daily until treatment completion and at two late follow up visits. During CHMI, daily monitoring of parasitaemia will be done to initiate treatment and reduce potential symptoms of malaria. Phlebotomies will be performed by qualified nurses and physicians and sites of sampling will be frequently inspected. For blood sampling in short intervals an intravenous catheter may be used. No more than 500 mL of blood (the equivalent of one blood donation) will be sampled over the study period.
*Other burden and risks*: Wild *Anopheles* mosquitoes may be infected by a participant. The risk is well below the background (imported infections from endemic areas) as gametocytes will circulate only shortly and at very low numbers. In addition, there are very few competent *Anopheles* species in the study area.
*Benefits*: There is no direct benefit for study subjects from participation in the trial. Subjects may indirectly benefit from general medical evaluation and health screening procedures. The group benefit of the study is mainly for people at risk for malaria; mostly in malaria-endemic regions but also travellers as RaViCHMI1 is intended to accelerate development of antimalarial interventions.

### 1. INTRODUCTION AND RATIONALE

*Plasmodium vivax* is the second-most important malaria parasite (following *P. falciparum*) and has a significant, although difficult to measure, disease burden [1], particularly in South and Southeast Asia as well as in South America. In parts of Africa, *P. vivax* infection also results in a significant disease burden. The World Health Organization has therefore included the development of vaccines to prevent *P. vivax* malaria and transmission into its updated Malaria Vaccine Technology Roadmap [2]. In order to assess the efficacy of malaria vaccine candidates, controlled human malaria infections (CHMI) play a pivotal role. This validated tool allows down-selection of vaccine candidates very early in clinical development based on the most relevant criterion – protective efficacy – with a minimal number of healthy and non-vulnerable participants. The gain in statistical power is achieved through highly controlled experimental conditions with regards to the sample (e.g. similar background immunity, age, and health status) and an inoculum that consistently leads to infection with similar kinetics. CHMI with *P. falciparum* is well established in the development pipeline for pre-erythrocytic, asexual blood stage and, since recently, also transmission-blocking vaccine candidates [3–6].

Today, P. *falciparum* CHMI is one of the most-used experimental models for vaccine development [7]. However, in the past, deliberate infection of humans with *P. vivax* was much more common. It was used extensively in the first half of the 20$^{th}$ century for the treatment of late-stage syphilis (i.e. malariotherapy). It was considered a standard therapy and was experimentally used for other conditions; some until the second half of the 20$^{th}$ century. It was one of the first immune-therapies (the malaria-induced systemic inflammatory reaction was the therapeutic principle), had an estimated efficacy around 50% and was used in tens of thousands of patients [8]. Until today, *P. vivax* CHMI plays a role in testing interventions, although the lack of standardised procedures and isolates as well as the scarcity of new interventions for *P. vivax* and difficulties in accessing well-defined challenge agents make them an underused technology.

Technologies of *P. falciparum* CHMI cannot be translated directly to *P. vivax* CHMI, because the biology of *P. vivax* is different from *P. falciparum*: 1) *P. vivax* parasites can reside in the liver for long time intervals (as so-called hypnozoites) and one infection event can lead to multiple episodes (in fact, most *P. vivax* malaria episodes are caused by relapses due to aroused hypnozoites), 2) *P. vivax* asexual blood stages cannot be kept reproducibly in continuous culture. This is due to its preference for invading reticulocytes and the unavailability of suitable culture conditions. In addition, cytoadhesive properties of *P. vivax* parasites are different from *P. falciparum*, which impacts the partitioning of the parasites between tissue and circulation as well as the time until a human infection becomes infectious to mosquitoes. In *P. vivax*, the infectious stage of the parasite circulates much earlier following inoculation compared to *P. falciparum*.

Recently, a protocol for asexual blood stage CHMI with PvW1, a *P. vivax* clone from Thailand has been established [4]. It uses a leuko-depleted, cryopreserved blood sample from a donor, who has blood group 0, is Rhesus and Kell antigen negative and whose blood has been safety screened. The inoculum has been tested in 37 participants so far. For PvW1, a high-quality genome assembly is available (GCA_914969965 at European Nucleotide Archive). A standardised blood stage inoculum with a *P. vivax* clone is a significant advance in malaria research. Besides PvW1, HMP013, one other *P. vivax* blood stage inoculum has been manufactured and used for CHMI since 2014. It has been cryopreserved directly from the blood of a traveller returning from India [6]. Blood stage inocula from malaria parasites can be used to assess the efficacy of drugs and vaccines targeted at the asexual and sexual blood stages (i.e. against the symptomatic and the transmission stage of malaria). To assess interventions that target the sporozoite and hepatic stages of the cycle, infected mosquitoes are required. As *P. vivax* cannot be maintained in continuous culture, blood from infected humans or animal models is required to feed mosquitoes. In some recent studies, infection of mosquitoes during CHMI was explored but no consistent protocol for transmission was developed [4,6]. The main obstacle for such a protocol is that *P. vivax* infection can lead to symptoms, which limit the time and parasitaemia easily tolerated by the participants. In times of malariotherapy those symptoms were the aim of *P. vivax* infections, and more systematic studies on transmission to mosquitoes were possible [9]. In RaViCHMI1, parasitaemia and symptoms will be kept to a minimum by using antimalarials that do not decrease gametocyte viability and infectivity (“gametocyte-sparing” treatment) as well as *ex vivo* concentration techniques before feeding of mosquitoes. Both approaches have been successfully used in CHMI studies before.

In RaViCHMI1, a protocol for reproducibly infecting laboratory-reared mosquitoes following inoculation of healthy, adult, malaria-naïve participants with asexual blood stage parasites of the PvW1 clone of *P. vivax* will be established. This model may be used to 1) assess interventions that inhibit transmission to mosquitoes (e.g. transmission-blocking vaccines) and to 2) produce infected mosquitoes to assess interventions targeting the sporozoite and hepatic stages of the parasite life cycle.

#### 2. OBJECTIVES

Primary objective:

- Develop a protocol to reproducibly infect mosquitoes with blood from human study participants undergoing asexual blood stage CHMI with the *P. vivax* clone PvW1

Secondary objectives:

Maximize mosquito infection rate with PvW1 by

- using *ex vivo* gametocyte concentration techniques, and
- antimalarial treatment that minimizes malaria-associated symptoms and adverse events due to asexual parasitaemia while not affecting gametocyte viability

Exploratory Objectives:

- Phenotype the immune response to asexual blood stage CHMI with *P. vivax*
- Assess *ex vivo* infectivity and development of *P. vivax* sporozoites in hepatocytes
- Measure the effect of *P. vivax* CHMI on sleep and metabolism
- Associate serum concentration of gametocyte-sparing antimalarials with infectivity and parasite stage distribution and gametocyte sex ratio

#### 3. STUDY DESIGN

RaViCHMI1 is a single centre, sequential, non-controlled, non-randomized, open-label study to establish and optimise a *P. vivax* human-to-mosquito transmission model. Mosquitoes will be infected following CHMI with a *P. vivax* blood-stage inoculum of the clone PvW1. In subsequent studies this CHMI model will be used to assess interventions that are designed to reduce *P. vivax* transmission from human gametocyte carriers to mosquitoes. It can also be used to assess interventions against the asexual blood stage of *P. vivax* malaria. In addition, *P. vivax*-infected mosquitoes will be produced. These infected mosquitoes may be used for *P. vivax* sporozoite human challenges to assess interventions against sporozoites and hepatic stage parasites including hypnozoites. In addition, sporozoites will be isolated from mosquito salivary glands to assess infectivity *ex vivo* and discover improved vaccine candidates.

RaViCHMI1 will be conducted at the Radboudumc in Nijmegen, The Netherlands. The anticipated time frame is Q2 2024 to Q2 2025. The study will recruit healthy, malaria-naïve adults in three groups of 4 participants (total n=12). The three groups will be recruited sequentially. All participants will undergo CHMI (i.e. no control group). Allocation to the groups will be by sequence.

All participants will be inoculated intravenously with the cryopreserved *P. vivax* W1 (PvW1) blood-stage parasite stabilate, using a previously established dose of the inoculum. Participants will be screened to minimise their risk due to participation in the study. Inoculation (Day+0) will be done at one time point for each group. The day before inoculation (Day-1) in- and exclusion criteria will be re-reviewed, and participants will have the opportunity to ask further questions. The day following inoculation (Day+1), participants will be seen by the clinical team to solicit adverse events and obtain a sample for immunological investigations.

From Day+2 to Day+4 participants will be called daily to check for potential Adverse Events (AEs). Malaria symptoms are expected to occur from about Day+10 onwards. From Day+5 onwards, participants will come daily to the study site. Here, vital signs will be recorded, AEs solicited, and a blood sample taken for the assessment of total parasitaemia (asexual plus gametocytes) by polymerase chain reaction (PCR). Hematologic and biochemical measurements will be done at pre-specified days (Appendix 1: Schedule of Procedure) and if deemed necessary by the investigator.

Beginning with Day+10, daily thick blood smear (TBS) will be done and asexual blood stage parasitaemia, gametocyte number as well sex ratio will be quantified by PCR. Based on data from 37 participants included in previous trials who were inoculated with PvW1, it is expected that TBS will become positive around Day+14. TBS will be used as the gold-standard diagnostic test.

Participants will be treated with a gametocyte-sparing antimalarial (Figure 1) when I) parasitaemia is >10 parasites/µL, or II) they have any parasitaemia and a body temperature >38.5°C and/or symptoms of malaria that significantly interfere with their daily life (e.g. severe symptoms or signs related to infection, including, but not limited to, rigors, subjective feverishness, sweats, headache, myalgia, arthralgia, nausea or vomiting).

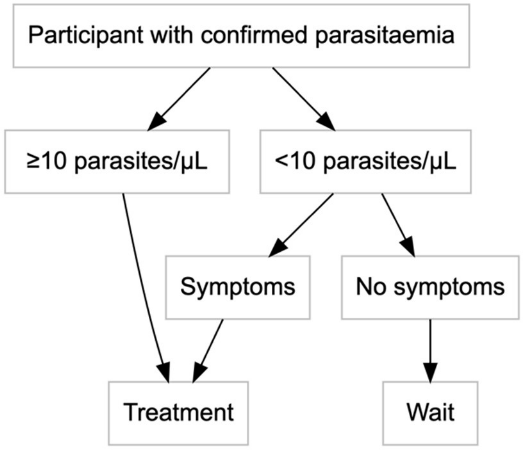

*Figure 1: Treatment algorithm. From Day+5 to Day+9, parasitaemia is measured by PCR (confirmation of successful infection). The threshold of 10 parasites/µL is determined by TBS, with PCR as backup.*

Two gametocyte-sparing treatments will be used: mepacrine (Group M) and piperaquine (Group P). Subsequently, a third group (Group O) will be recruited to optimize the gametocyte-sparing treatment by selection of the drug and dose. Group O is conditional on the results of Group M and Group P. The sequence of Groups M and P is not predefined, whereas Group O follows the other two groups. The treatment used in Group O will be chosen following review of Group M and Group P data.

Following the decision to treat, mepacrine will be used at a dose of 100 mg every 8 hours and 100 mg per day the following 3 days (Group M). Mepacrine, 100 mg per day, is a suppressive dose that was used for the prevention of *P. vivax* relapse before 8-aminoquinolines were available [10]. The volume of distribution of mepacrine is very large, therefore a loading dose is required to achieve high enough drug levels early on [11]. In Group P, piperaquine will be used as a single dose of 480 mg. This regimen has been successful in *P. falciparum* transmission studies [3].

Human-to-mosquito transmission will be measured by mosquito feeding assays (Direct Membrane Feeding Assay – DMFA) using the participants' blood and concentrated gametocytes as well as by feeding on the skin of the participants.

All mosquito feeding assays will be done between Day+10 and Day+26. The timepoints of the mosquito feeding assays will be adapted based on the results that we will gather from the different cohorts. The sequential groups will allow us to focus towards the main aim of the study. We will optimize the procedures in three steps.

- First Group: Measure transmission kinetics and identify best gametocyte enrichment method
- Second Group: Confirm transmission peak and best gametocyte enrichment method
- Group O: Confirm best transmission time and method and produce infected mosquitoes.

While adapting the timepoints and procedures based on the results of the preceding group, we will adhere to the following in all groups: there will be no more than four skin feeding assays and we will not exceed the maximum amount of bloodwithdrawal. The number of blood draws will reduce with the sequence of groups, while the amount per sample may rise.

Based on experimental evidence, mepacrine and piperaquine are expected to kill asexual parasites whilst leaving gametocytes intact. When using gametocyte-sparing drugs that act against asexual parasites such as mepacrine or piperaquine, gametocytaemia will decrease with a lag of 2-3 days [9], because the multiplication cycle is interrupted and no newly sexual-development committed ring stage parasites are produced. In previous studies using the same inoculum, treatment was initiated around Day+15. The third group (Group O) will be used to increase power for the estimate of the mosquito infection rate of the best treatment. Treatment regimen may be altered to allow for further optimization of the treatment strategy.

All participants will receive an end-of-study treatment with either atovaquone-proguanil (first line) or artemether-lumefantrine (second line). End-of-study treatment will be given:

- On Day+24 to participants who develop parasitaemia <10 parasites/µL and no symptoms
- On Day+24 to participants who develop neither detectable parasitaemia nor symptoms
- On Day+24 to participants receiving gametocyte-sparing treatment with mepacrine or piperaquine who tolerate symptoms (if any), do not develop Grade 3 AEs or Serious Adverse Events (SAEs) and, have a parasitaemia <500 parasites/µL.
- Within 24 hours to participants receiving gametocyte-sparing treatment who experience related SAEs, Grade 3 AEs related to gametocyte sparing medication or those who develop a parasitemia of more than 500 parasites/microliter on two subsequent days; in case of Grade 3 malaria-related symptoms, the decision to treat with end-of-study treatment may

be delayed for up to 48 hours, given that the participant agrees and symptoms decrease in severity over that period.

In all cases, the investigator may initiate treatment earlier if deemed necessary based on parasitological or clinical judgement. In all cases, clearance of parasitaemia will be verified following completion of end-of-study treatment.

**Group M**: Group M participants will start a treatment regime of orally administered mepacrine (mepacrine dihydrochloride, 100 mg every 8 hours on the first day, followed by 100 mg once daily for 3 days) upon reaching the symptom threshold in combination with any parasitaemia (body temperature >38.5°C and/or symptoms of malaria that significantly interfere with daily life), or upon developing a parasitaemia >10 parasites per µL. Mepacrine, 100 mg three times a day, is a commonly used treatment for treatment-refractory giardiasis. In the past, doses of up to 1 g per day have been used in malaria treatment [11]. Doses as little as 50 mg per day have been reported to suppress asexual *P. vivax* replication but leave patients infectious to mosquitoes for a further ~3 days [9].

**Group P**: Group P participants will receive 480 mg piperaquine orally, which has been shown to clear asexual parasitaemia in a *P. falciparum* CHMI whilst leaving gametocytes to circulate that are infectious to mosquitoes. Treatment will be initiated as described for mepacrine. There is no systematic data on piperaquine monotherapy of *P. vivax* infections. In combination with dihydroartemisinin it has been reported to be highly efficacious and providing a significant suppressive effect [12].

**Group O**: The treatment regime for Group O will be finalised following review of Group M and P data. The purpose thereof is either to increase power of Group M or P or to further optimise the *P. vivax* CHMI transmission model in terms of transmissibility versus safety/tolerability.

**Selection of the drug in Group O:**

- The treatment that results in peak infection rates of mosquitoes >80% and is safe and well tolerated is selected
- In case mosquito infection in either Group M or P is successful, the treatment with higher mosquito infection rate and higher salivary gland sporozoite count will be used
- In case that peak infection rates are >80% in both groups and result in similar salivary sporozoite counts, the better tolerated treatment will be chosen
- In case that none of the treatment works the dose of mepacrine or piperaquine will be adapted.

**Treatment adaptions in case that both gametocyte-sparing treatments do not meet criteria on infection or tolerability:**

- Peak infection rate is <80%:
  - Update treatment criterion to >100 parasites/µL and/or any parasitaemia with symptoms of malaria that significantly interfere with daily life
  - Reduce dose of mepacrine to 50 mg every 8 hours on first day, followed by 50 mg per day for 3 days
  - Reduce piperaquine dose to 320 mg
- CHMI not well tolerated:
  - Update treatment criterion to: >1 parasite/µL and/or symptoms of malaria that significantly interfere with daily life
  - Incomplete mepacrine suppression of parasite multiplication – increase loading dose to 200 mg every 6 hours on the first day (total 1 g), followed by 100 mg daily
  - Incomplete piperaquine suppression of parasite multiplication – increase in piperaquine dose to 960 mg

**Serum concentration of gametocyte-sparing antimalarials**

Serum concentration of gametocyte-sparing antimalarials will be assessed. Blood will be sampled at pre-defined time points to measure plasma and erythrocyte drug concentrations to assess the effect on *P. vivax* blood-stage parasitaemia, stage-distribution, gametocyte count and sex-ratio. For that purpose, asexual and sexual stage parasites will be analysed by molecular methods.

<u>Sampling schedule Group M</u>

Before dosing, 1-, 2-, 3- and 7-days post-dosing; Late follow up visits: Day+26, Day+35 and Day+49.

<u>Sampling schedule Group P</u>

Before dosing, 3-, 6-, 8- and 24-hours and 7-days post-dosing. Late follow-up visits: Day+26 and Day+35. Sampling timepoints after 3 hours following administration of gametocyte-sparing antimalarial and before 24 hours post-dosing might be delayed by up to 5 hours.

The sampling schedule for Group O will follow either Group M or Group P, depending on the used gametocyte-sparing antimalarial.

**Use of wearables and questionnaires to assess metabolism and sleep**

Sleep and metabolism will be measured for the duration of the study, to allow for detection of potential differences in behavioural as well sleep patterns at baseline and during CHMI and their effect on immunological and metabolic responses. Measurements will be taken using wearable devices, capable of measuring rest-activity rhythms and sleep patterns, vital signs, and temperature. The devices will be provided for the duration of the study and used by the participants at home. In addition, sleep measurements will be validated using a portable full EEG measurement device that can be used at home. Additionally, diaries and standardized questionnaires recording sleep pattern and dream-related questions will be used.

**Immunology**

Peripheral blood mononuclear cells (PBMC) will be collected at baseline, Day+1 and on the day of first dose of an antimalarial. Cells will be cryopreserved and phenotyped using high-dimensional single cell measurement by cytometry, single cell sequencing and cell metabolism profiling. Serum will be collected at screening, Day+35 and Day+49 for serologic profiling against plasmodial and cross-reacting antigens, hormones, metabolites and cytokines.

***Ex vivo* infection studies**

Salivary gland sporozoites will be collected from *Anopheles* mosquitos fed by gametocyte enriched blood. Sporozoites will be analysed on the transcriptome, proteome and epigenome level and used to infect cultured primary human or macaque hepatocytes. Infected hepatocytes will then be analysed on the phenotypic and molecular level including highly dimensional methods.

### 4. STUDY POPULATION

#### 4.1. Population

Healthy, malaria-naïve adult male and female volunteers aged 18-45 years will be recruited and 12 participants will be enrolled into the study. In addition, up to 3 reserve participants (1 reserve participant per group) will be recruited to act as a replacement in case a volunteer drops out before CHMI. The investigator will ensure that all participants being considered for the study meet the eligibility criteria. A relevant record of the eligibility criteria assessment will be stored with the source documentation at the study site.

#### 4.2. Inclusion criteria

To be eligible to participate in this study, a potential participant must meet all of the following criteria:

- Written informed consent
- Successful completion of study quiz
- Healthy, malaria-naïve volunteer aged 18-45 years
- General good health based on history and clinical examination
- Willing to remain within 2 hours travelling distance from the study centre for the duration of CHMI (inoculation to end-of-study treatment)
- Reachable 24/7 by mobile phone during the CHMI period
- Women of childbearing potential: must agree to practise continuous effective contraception for the duration of the study
- Agree to refrain from blood donation throughout the study period
- Agree to refrain from intensive physical exercise (disproportionate to the subject's usual daily activity or exercise routine) during the CHMI period until one week after completion of treatment
- Agree to their general practitioner (GP) being informed about participation in the study and agree to sign a form to request the release by their GP, and medical specialist when necessary, of any relevant medical information concerning possible contraindications for participation in the study to the investigator
- Able and willing (in the investigator's opinion) to comply with all trial requirements

#### 4.3. Exclusion criteria

A potential participant who meets any of the following criteria will be excluded from participation in this study:

- History of clinical malaria (any species)
- Red blood cells negative for the Duffy antigen/chemokine receptor (DARC)
- For women: red blood cells negative for Rhesus antigen c (little c)
- Known or suspected haemolytic disease or presence of hemoglobinopathies
- Use of systemic antibiotics with known antimalarial activity within 30 days of study enrolment (e.g. trimethoprim-sulfamethoxazole, doxycycline, tetracycline, clindamycin, erythromycin, fluoroquinolones, or azithromycin)
- Travel to a region where malaria prophylaxis is recommended within the preceding three months or planned travel during the study period
- Any clinically significant abnormal finding on clinical examination or laboratory screening
- Any confirmed or suspected immunosuppressive or immunodeficient state, including asplenia; recurrent, severe infections and chronic (more than 14 days) immunosuppressant medication within the past 6 months (inhaled and topical steroids are allowed).
- Positive urine toxicology test for cannabis, cocaine, or amphetamines at inclusion.
- Screening tests positive for Human Immunodeficiency Virus (HIV), active Hepatitis B Virus (HBV), or Hepatitis C Virus (HCV).
- Screening test positive for West Nile virus (WNV) IgM antibodies
- Receipt of any investigational or non-registered product (drug or vaccine) in the 28 days preceding enrolment or during the study period.
- Participation in any other clinical study in the 28 days prior to the start of the study or during the study period
- Previous participation in a malaria vaccine trial
- Administration of immunoglobulins and/or any blood products within the three months prior to the start of the study or planned administration during the study period
- Pregnancy, lactation or intention to become pregnant during the study period
- History of drug or alcohol abuse interfering with normal functioning in the five years preceding enrolment
- Falling in moderate risk or higher categories for fatal or non-fatal cardiovascular events within 5 years (>5%) determined by non-invasive criteria for cardiac risk [13]
- A positive family history of cardiac events in 1st or 2nd degree relatives <50 years old

- Abnormal electrocardiogram on screening: pathologic Q wave and significant ST-T wave changes, left ventricular hypertrophy, clinically significant arrhythmias, left bundle branch block, secondary or tertiary A-V heart block
- A QT/QTcB interval >450 ms
- Current use of medications known to cause prolongation of the QT interval
- Contra-indications to the use of the antimalarial medications used in this study
- History of clotting disorder
- History of seizure, except for sporadic febrile convulsions in childhood
- History of serious psychiatric condition that may affect participation in the study (including but not restricted to organic, including symptomatic, mental disorders [ICD-10 code: F00-F09], schizophrenia, schizotypal and delusional disorders [F20-F29], mood (affective) disorders [F30-F39], mental retardation [F70-F79], disorders of psychological development [F80-F89] or any other psychiatric condition that required hospitalization or psychiatric treatment over an extended period).
- History of cancer, except cervical carcinoma in situ
- Being an employee or student of the Department of Medical Microbiology of the Radboudumc, or a person otherwise related to the investigator other than a professional relationship for clinical study purpose only
- Any other condition or situation that would, in the opinion of the investigator, place the participant at an unacceptable risk of injury, affect the ability of the participant to participate in the study, or impair interpretation of the study data

##### 4.4. Power considerations

A total of 12 participants will be enrolled in three groups of 4 participants. As 37/37 participants that have been inoculated with PvW1 became positive, the 95% binomial confidence interval of parasitaemia is 90.5–100%. Given a conservative assumption that infection success is at the lower side of the interval, there is still <1% probability that 4/4 CHMI attempts are not successful. In previous studies, about 50% of the volunteers do not develop at least moderate (Grade 2) symptoms and fever at a parasitaemia of ≥10 per µL. Consequently, the sample size to be able to perform with 90% probability a delayed treatment in at least one participant at a parasitaemia >10 parasites per µL, which increases the probability of successful transmission, is 4. In the event that none of the participants achieve a peak parasitaemia of ≥10 per µL in the first or second cohort and in case that the best feeding results are below 10% infected mosquitoes, the sample size may be recalculated up to two times based on the evidence accumulated following completion of the respective groups.

Continuation of the study will depend on ethical and regulatory approval of a major amendment of the dossier that contains the updated sample size.

### 5. TREATMENT OF SUBJECTS

All participants will undergo CHMI with *P. vivax* using a blood inoculum containing the clone PvW1. The primary objective of the study is to establish and optimise a CHMI model for transmission of *P. vivax* from infected humans to *Anopheles* mosquitoes. Clearing asexual blood-stage parasites while sparing gametocytes to infect mosquitoes shall be achieved by administering the antimalarials mepacrine or piperaquine. End-of-study treatment will be atovaquone-proguanil (first line) or artemether lumefantrine (second line).

#### 5.1. Investigational product/treatment

Not applicable

#### 5.2. Use of co-intervention

**Inoculum**

All participants will undergo a controlled human *P. vivax* infection using a previously characterised asexual blood stage inoculum containing PvW1, a *P. vivax clone* originally isolated in Thailand [4]. Participants will be inoculated intravenously with a standard dose of leukocyte-depleted, cryopreserved blood sample from a universal blood donor, who was infected with PvW1. Inoculation will be done according to an established standard operating procedure used in previous trials. The inoculum contains approximately 2,600 blood-stage *P. vivax* parasites in about 50µL of erythrocyte pellet.

**Gametocyte-sparing antimalarial**

Two different registered anti-malarial drugs will be used to treat asexual blood stage parasites without affecting *P. vivax* gametocytes:

*Mepacrine*: participants in Group M will be treated with mepacrine when parasitaemia is ≥10 parasites/µL or when any parasitaemia and significant malaria-related symptoms occur (Figure 1).

*Piperaquine*: participants in Group P will be treated with 480 mg piperaquine, a dose that has been successfully used in a *P. falciparum* transmission model. Treatment initiation is as for mepacrine.

The treatment regime in Group O will be selected following review of Group M and P data, with the purpose to select a regimen for future studies and further optimise the *P. vivax* CHMI transmission model.

**End-of-study treatment**

All volunteers who developed parasitaemia will receive an end-of-study treatment. In line with Dutch national parasitology guidelines the treatment will consist of:

- First line: atovaquone-proguanil (4 tablets of 250-100 mg orally once a day for 3 days together with fatty food or a milky drink)
- Second line: artemether-lumefantrine (80/480mg orally at 0, 8, 24, 36, 48 and 60 hours; together with fatty food or a milky drink)
- In case first and second line cannot be used: any other anti-malarial regimen registered for the treatment of *P. vivax*

Since controlled human *P. vivax* infections will be induced in all groups of participants by inoculation of a blood-stage inoculum, additional (radical) treatment with primaquine to clear hypnozoites is not necessary.

**Contraceptives**

Female participants of child-bearing potential must use one or more adequate contraceptive methods throughout the study if sexually active. Acceptable forms of contraception include: use of oral, injected or implanted hormonal contraceptives; intrauterine device or intrauterine system; barrier methods (condoms or diaphragm with additional spermicide); male partner's sterilisation (with appropriate post-vasectomy documentation of absence of sperm in the ejaculate). No contraceptive methods are required in case of true abstinence, when this is in line with the preferred and usual lifestyle of the subject, but periodic abstinence (e.g., calendar, ovulation, symptothermal, post-ovulation methods) and withdrawal are not acceptable methods of contraception. Note that female participants using (systemic) hormonal contraceptives and that are treated with artemether-lumefantrine are advised to use an additional non-hormonal contraceptive method for at least 1 month after treatment.

**Other drugs with antimalarial activity**

Throughout the study, participants should avoid taking antimalarials other than those prescribed by the clinical team, as well as antibiotics and other drugs with potential antimalarial properties, such as tetracyclines, macrolides/lincosamides or co-trimoxazole. If in doubt participants or their prescribing physician should discuss the relative benefits and risks with the clinical team.

**Drug-drug interactions**

Participants receiving piperaquine or mepacrine should avoid use of concomitant drugs that prolong the QTc-interval. In addition, drugs with strong CYP3A4-inhibitory or -stimulatory activity (note that also St. John's wort belongs to this class), drugs that are metabolised by CYP3A4, CYP2C19 (e.g., citalopram) or CYP2E1 and have a narrow therapeutic window shall be avoided.

Participants receiving artemether-lumefantrine should avoid concomitant use of drugs that prolong the QTc-interval. Female participants using systemic hormonal contraceptives are advised to use an additional non-hormonal contraceptive method for at least 1 month after taking artemether-lumefantrine. In addition, interactions can occur with drugs with strong CYP3A4-inhibitory or -stimulatory activity, drugs that are metabolised by CYP2D6, as well as grapefruit juice.

#### 5.3. Escape medication

Escape medication during CHMI consists of end-of-study treatment. All participants will receive an end-of-study treatment (see above) with either atovaquone-proguanil (first line) or artemether-lumefantrine (second line). If oral medication cannot be taken by the participant (e.g. because of vomiting), parenteral artesunate will be used as a first line treatment until it can be switched back to an oral regimen.

In all cases, clearance of parasitaemia will be verified following completion of end-of-study treatment.

End-of-study treatment may be initiated earlier than scheduled in any given participant if deemed necessary by the investigator based on clinical judgement or because the participant retracts consent in participation.

Symptomatic treatment for malaria-related symptoms will be provided to participants, as appropriate. This includes the use of paracetamol, NSAIDs and anti-nausea medication.

### 6. INVESTIGATIONAL PRODUCT

Not applicable

### 7. NON-INVESTIGATIONAL PRODUCT

#### 7.1. Name and description of non-investigational product(s)

##### 7.1.1. Plasmodium vivax W1 challenge inoculum

The *P. vivax* W1 challenge inoculum PvW1 is a *P. vivax* clone from Thailand. It is a leukocyte-depleted, cryopreserved blood sample from a donor, who has blood group 0, is Rhesus and Kell antigen negative and whose blood has been screened for infections and microbial contamination. For PvW1, a high-quality genome assembly is available (GCA_914969965 at European Nucleotide Archive).

##### 7.1.2. Antimalarials

*Mepacrine*: Mepacrine will be used as a gametocyte-sparing treatment in Group M and potentially Group O. Mepacrine, also known as quinacrine or atebrine was the first synthetic antimalarial in wide use. Nowadays, it is still used as a treatment for therapy-resistant giardiasis and has been experimentally used for various infectious, autoimmune, and neoplastic diseases. In the dose used in this trial, mepacrine is generally well tolerated. Possible adverse effects are: yellow discoloration of the skin and urine during long-term treatment or with large doses, blue or black discolouration of the roof of the mouth, nails and eyes (this resolves on discontinuation of the drug), dizziness, particularly after sitting or lying down. Other possible side effects include nausea, vomiting, gastrointestinal upset, headache, skin rashes (occasionally severe), and changes in mood or behaviour. Fits may occur with overdosing. Alterations in blood count and liver function can occur but are rare [14].

*Piperaquine*: Piperaquine will be used as a gametocyte-sparing treatment in Group P and potentially Group O. It will be administered orally at a dose of 480 mg. Dosing is based on experience with gametocyte-sparing treatments to set up a transmission model at Radboudumc using *P. falciparum* CHMI [3]. When used for the treatment of malaria in combination with dihydroartemisinin, common adverse reactions (≥10%) include tachycardia, QT-interval prolongation, headache, fever, asthenia, anaemia.

*Atovaquone-proguanil*: atovaquone-proguanil is a marketed and recommended treatment for *P. falciparum* and *P. vivax* malaria. It will be used as first-line end-of-study treatment. Common adverse reactions (≥10%) in adults associated with atovaquone-proguanil treatment include headache, nausea and vomiting, diarrhoea, and abdominal pain. Unlike artemether-lumefantrine and chloroquine, atovaquone-proguanil does not cause QT-interval prolongation.

*Artemether-lumefantrine*: artemether-lumefantrine is a marketed and recommended treatment for *P. falciparum* and *P. vivax* malaria. It will be used as second-line end-of-study treatment. Common adverse reactions (≥10%) include headache, dizziness, heart palpitations, nausea and vomiting, abdominal pain, fatigue, loss of appetite, asthenia, myalgia, and arthralgia. Use of artemether-lumefantrine has been associated with QT-interval prolongation.

*Use of an unauthorized auxiliary medicinal product:* Mepacrine is an unauthorized medicinal product in the European Union. Nowadays, it is used on the basis of patient-specific prescriptions to treat parasitic diseases (e.g. therapy-refractory giardiasis) and immune disorders (e.g. discoid lupus erythematosus, subacute cutaneous lupus erythematosus, erythema multiforme, sarcoidosis and dermatomyositis). Patient information leaflets are available but no SmPC. Mepacrine is on the US FDA 503b bulks list and marketed in countries outside the EU, which do not use a SmPC. In the era when mepacine was heavily used (i.e. around the 2nd world war it was used in millions of doses), the current system of documentation was also not in place. The product that will be used is produced according to cGMP for use in the study. The scientific reason for the use of mepacrine in RaViCHMI1 is based on an extensive review of the literature and own data from clinical trials *in P. vivax*-endemic regions. It emerged as the only product with experimental evidence that it acts against asexual but not sexual blood stage *P. vivax* parasites. The goal of the study is to infect mosquitoes while performing a CHMI that is well-tolerated for the participants. Mepacrine is the most promising candidate to achieve high infection rates in a safe and manner. As clinical experience is large and a compliant product is available, we argue that the use of the unauthorized product mepacrine is reasonable.

#### 7.2. Summary of findings from non-clinical and clinical studies

Deliberate infections with malaria parasites have been done since 1917, first with the intention to induce fever and treat patients with late-stage syphilis [8]; one of the first systematically used immunotherapies. Malariotherapy became a standard treatment, used in many thousands of patients worldwide. Later, controlled infections became a major tool in the development of new interventions. CHMI remains a powerful tool for clinical research. Radboudumc is among the leading centres for CHMI studies in the world and the only centre in Europe that continuously maintains the full lifecycle of the *P. falciparum* parasite in its natural host species.
The majority of modern CHMI studies (including all CHMI studies in Radboudumc) have used *P. falciparum*, either using sporozoite inoculation by mosquito bite or by parenteral inoculation of purified, cryopreserved sporozoites (PfSPZ), or by intravenous injection of asexual blood-stage parasites (also called induced blood stage malaria – IBSM). Although CHMI with *P. vivax* is not common, there has been a recent increase of interest that led to the development of more

standardized challenge models in Colombia, Australia, US and UK. The PvW1 inoculum that will be used in this study has been tested in 37 participants, so far.

#### 7.3. Summary of known and potential risks and benefits

##### 7.3.1. Risk assessment / structured risk analysis

Risks of the study can be divided into risks associated with the inoculum, *P. vivax* malaria, antimalarial treatment, mosquito bites, and transmission of malaria. In the following, a structured risk analysis is provided followed by mitigation strategies that will be used in this study.

**Risks associated with the inoculum**

The donor blood is Blood Group O, Rhesus (Rh) and Kell antigen negative. In contrast to common procedures to obtain infected blood, the donor has been prospectively screened to be a universal blood donor and pass all criteria for blood donation before being infected. The risk of a transfusion reaction or development of antibodies to donor red blood cells is therefore minimal. The inoculum corresponds to approximately 50 µL of packed red cells. Furthermore, the inoculum is leukocyte depleted and tested for microbial contamination. All procedures related to producing the blood inoculum were done under strict quality assurance. The inoculum has been used safely in 37 malaria-naïve participants in the UK, so far.

*Risk mitigation*

Following inoculation, the participants will stay at the clinical ward and are monitored for at least one hour. Vital signs will be taken before inoculation, and 15 minutes and 1 hour thereafter. If needed, more and more intense monitoring will be done. A trained physician will be present during the monitoring period. Inoculation by intravenous injection will be performed by a trained member of the clinical team. The clinical team is proficient in advanced adult life support and the treatment of anaphylactic reactions. A kit to treat anaphylactic reactions is present at bedside. In case of severe reactions, Radboudumc is fully equipped to manage anaphylaxis and any other medical emergency including re-animation and intensive care.

**Risks associated with *P. vivax* malaria**

The main burden of *P. vivax* malaria are symptoms of systemic inflammation, such as fatigue, fever, headache, and myalgia. *P. vivax* infection is commonly characterized as “benign tertian malaria” because, in contrast to *P. falciparum* malaria, the risk of complications in healthy adults with an acute acquired infection is very low. Nevertheless, there is a significant global disease burden including complications and death, which are mostly associated with chronic and relapsing infections, pregnancy, and co-morbidities [1]. In previous *P. vivax* CHMI studies few participants received intravenous fluids because of nausea and vomiting [15].

There is no risk of *P. vivax* relapse because a blood inoculum is used (relapse is caused by liver stage hypnozoites). The antimalarial treatment is curative, therefore no complications of chronic or recurrent malaria may occur.

*Risk mitigation*

It will be ensured that study participants understand all risks associated with malaria. As part of the inclusion procedure, volunteers must pass a written test that shows that they actively understand the study procedures and the associated risks. Contact details of at least one close relative or friend of the study participant who is informed about the whereabouts of the volunteer will be collected, to be able to locate the study participant even when they are not compliant. Volunteers will be counselled that should they fail to return for treatment after having been infected with *P. vivax* they could become very unwell and experience complications.

In the unlikely event that a volunteer after being inoculated and before completion of an appropriate course of antimalarial therapy should fail to show up for a scheduled clinical visit or be unreachable by telephone or other means, all efforts will be made to locate the participant. The following resources may be used in an effort to locate the participant: the local police department, local accident and emergency departments and local, national and international media. While all parties will aim to preserve the participant's confidentiality, if necessary, details of the participant's identity (including photos) and participation in the study may be passed to authorities and the media in order to help locate the missing individual. Volunteers will be informed of this possibility during screening.

Participants will be followed up closely following CHMI, and treatment will be started at a conservative parasitaemia threshold (>10 parasites per µL) or if significant symptoms occur in combination with any parasitaemia.

**Risks associated with antimalarial treatment**

Participants will receive gametocyte-sparing antimalarial treatment with piperaquine or mepacrine as well as end-of-study treatment with atovaquone-proguanil or artemether-lumefantrine. All antimalarials used in the study have been given in millions of doses and their safety profile is well described. When given for the treatment of malaria common adverse effects of all antimalarials that will be used include headache, gastrointestinal symptoms, fever, and fatigue. A main risk in antimalarial treatment is non-adherence to the regimen.

Piperaquine, mepacrine and lumefantrine can prolong the QT interval. High doses or prolonged treatment with mepacrine (months or years), which can accumulate, can lead to skin coloration and can have psychological effects.

*Risk mitigation*

To ensure proper intake of the drugs, antimalarial treatment will be under observation by the clinical team. Where possible at the study site. Single doses may be taken at home with confirmation by phone or video. Atovaquone-proguanil and artemether-lumefantrine will be given with milk or fatty food to increase absorbance. In case that a volunteer vomits the medication within 30 minutes following drug administration, the full dose is repeated, between 30 and 60 minutes, half of the dose is given and after 60 minutes no re-dosing is done. When oral therapy is not possible, participants will be treated parenterally with artesunate until they tolerate oral medication. Mepacrine will be given at a low dose and over a short period of time (4 days). To mitigate cardiac risks, an electrocardiogram is part of screening. Potential participants with ECG abnormalities will be excluded. In case participants take co-medications, interactions on will be checked and monitored if required.

**Risks associated with phlebotomy**

There will be 27 scheduled visits with phlebotomies: at screening, before inoculation, daily until treatment completion and at two late follow up visits. During CHMI, daily monitoring of parasitaemia will be done to initiate treatment and reduce potential symptoms of malaria. Blood sampling can be associated with faintness, mild pain, bruising, irritation or redness at the site where blood was taken. In rare cases arteries or nervous tissue may be injured or a punctured vessel may occlude and induce inflammation of the surrounding tissue.

*Risk mitigation*

Phlebotomies will be performed by qualified nurses and physicians and sites of sampling will be frequently inspected. For blood sampling in short intervals an intravenous catheter may be used. No more than 500 mL of blood (the equivalent of one blood donation) will be sampled over the study period.

**Risks associated with parasite transmission**

Wild *Anopheles* mosquitoes may be infected by a participant and infected *Anopheles* mosquitoes may escape the insectary and infect non-participating individuals accidentally.

*Risk mitigation*

The risk of transmission is well below the background (imported infections from endemic areas [16]) as gametocytes will circulate only shortly and at very low numbers. In addition, there are very few competent *Anopheles* species in the study area.
Gametocyte numbers and infectivity are checked frequently during CHMI. Based on knowledge from previous studies [4,6], it is expected that infection rates will be very low, except for 1-2 days, where a small fraction highly competent vector (such as *Anopheles stephensi*) may get infected following skin feeding.
Besides strict procedures and training of the entomological team, a system of multiple doors and depressurization is present to prevent infected mosquitoes from accidentally escaping the insectary. In addition, only a fraction of infected mosquitoes will be allowed to be infected until sporozoites develop.

**General risk management**
All participants will be closely followed up by the clinical team for the duration of the study. PvW1 is a *P. vivax* clone that is sensitive to all marketed antimalarials that are recommended for treatment of *P. vivax*. This includes atovaquone-proguanil and artemether-lumefantrine.

##### 7.3.2. Benefits

There are no direct benefits for study participants from participation in the study. Study participants may indirectly benefit from general medical evaluation and health screening procedures including testing for HIV, hepatitis B, and hepatitis C. They will be informed about the results of the screening and if necessary, they will be referred to their primary physician where they will receive counselling and further medical attention. Study participants will receive a financial compensation which is reasonable and in line with Dutch common practice. The societal or group benefit of the study is mainly for people at risk for malaria; mostly in malaria-endemic regions but also travellers from regions without malaria as RaViCHMI1 is intended to accelerate development of antimalarial interventions and improve the impact of malaria control programs.

#### 7.4. Description and justification of route of administration and dosage

All study participants will be inoculated intravenously with 1:10 dilution of a cryopreserved PvW1 stabilate (corresponding to ~2,600 parasites and 50 µL packed red cells before freezing). This dose has been established to consistently infect human volunteers and allow up to 8 replication cycles before treatment initiation [4]. No other route has been tested.

##### 7.5. Dosages, dosage modifications and method of administration

The PvW1 challenge inoculum will be administered by intravenous injection into an indwelling intravenous cannula, followed by a saline flush. All procedures relating to thawing of the blood inoculum and preparation of the syringes will be performed in accordance with standard operating procedures. Participants will be observed for a minimum of one hour before discharge. Participants will receive the inoculum within a maximum of 4 hours of removal form the frozen storage.
All participants receiving the same dose of PvW1 parasites developed a blood-stage *P. vivax* infection. Therefore, dosage will not be modified in this study.

##### 7.6. Preparation and labelling of Non-Investigational Product

Preparation of the blood stage challenge product follows a standard operating procedure ensuring detailed documentation for each vial during the preparation process until administration to the study participant.
All other medication will be obtained directly through the Radboudumc pharmacy. Mepacrine and piperaquine will carry study specific labelling.

##### 7.7. Drug accountability

**Shipment, receipt and storage**

The non-investigational products will be shipped (and imported where applicable) to Radboudumc according to relevant national and international regulations. Shipments will be in accordance to the product's required specifications. Upon receipt at Radboudumc pharmacy the person in charge of product receipt will check that all specifications (e.g., cold chain) were maintained during shipment. In case of a problem, the clinical study monitor and the Sponsor must be alerted immediately. The acknowledgement of receipt will be dated and signed by the person in charge of product management and filed in the corresponding study file.
At Radboudumc, the PvW1 challenge inoculum will be stored in liquid nitrogen vapour phase in temperature-controlled liquid nitrogen containers with automated liquid nitrogen supply. Only authorized staff will have access to the storage area.
Malaria medication is stored in a dedicated on-site pharmacy room at the clinical ward.

**Accountability**

The person in charge of product management will maintain an inventory of all non-investigational products.

This person will also ensure the security of records and documents. In accordance with all applicable regulatory requirements, the person in charge of the product management at the site must keep up to date the following inventory records:

- Receipt of product at the clinical study site
- Product administered to each study participant
- Inventory of product at the clinical study site
- Log of any unused or expired product

These records should include dates, quantities, batch numbers, and period of use or expiration dates (if applicable). These records should also document adequately that:

- The study participants were provided the doses specified by the protocol
- All products provided are fully reconciled

These records will be monitored and verified by a monitor regularly, as part of the routine monitoring procedure as described in the Monitoring Plan.

**Return and destruction**

Unused or open products will be destroyed according to Radboudumc procedures.

### 8. METHODS

#### 8.1. Study parameters/endpoints

##### 8.1.1. Main study parameter/endpoint

Proportion of infected mosquitoes, measured as oocyst count of ≥1 oocyst following exposure to blood of participants with *P. vivax* parasitemia.

##### 8.1.2. Secondary study parameter/endpoint

Number and severity of adverse events related to CHMI between inoculation and completion of end-of-study treatment.

##### 8.1.3. Other study parameters

- Effect of time in days following inoculation and method of applying blood meal to mosquitoes: I) direct skin feeding, II) membrane feeding of whole blood, III) membrane feeding of gametocyte-enriched blood on mosquito infection rate.
- Mepacrine plasma and erythrocyte concentration
- Piperaquine plasma concentration
- *P. vivax* parasitaemia, gametocytaemia, sex and age distribution over time
- Cellular and humoral immune response pattern at baseline, during P. vivax parasitaemia and at reconvalescence
- Duration and pattern of sleep, sleep quality, rest-activity rhythms
- Dynamics of heart rate, body temperature, oxygen saturation, energy metabolism

#### 8.2. Randomisation, blinding and treatment allocation

No randomization or blinding will be done. All participants will receive the same inoculum at the same dose. Antimalarial treatment will be according to group allocation. Group allocation will be sequential. Group O follows Group P and M.

#### 8.3. Study procedures

**Mosquito feeding assays**

A reproducible CHMI transmission model requires enough mature gametocytes in the circulation of the study participants to ensure infection of mosquitoes through blood feeding. In this study three different mosquito feeding assays will be applied: I) direct skin feeding assay (DFA), II) DMFA and III)

DMFA with prior gametocyte enrichment.All feeding assays are performed with *Anopheles stephensi* mosquitoes reared at the Radboudumc insectary, that are collected in small cups or cages. Other mosquito species may be used for membrane feeding assays. All mosquitoes are laboratory strains and have been reared, for many years, under laboratory conditions and in climate controlled rooms. After the feedings the mosquitoes are given an extra blood meal with transfusion grade blood to speed up the parasite development in the mosquito.

For all assays unfed and partially unfed mosquitoes are removed from the cage after feeding. The infection status of mosquitoes fed with blood from the participants is assessed 6 to 9 days after the blood meal. The established oocysts in the midgut will be quantified using microscopy and following a standardized protocol [17]. Molecular methods may be used to complement findings or to increase throughput. A sample of the mosquitoes may be dissected up to 20 days following feed to assess development into sporozoite.

<u>Direct skin feeding</u>

For DFA study participants are instructed to place a forearm on a netted cup. Up to fifty mosquitoes are allowed to ingest a blood meal for 10 minutes on the volunteers exposed skin.

<u>Direct membrane feeding and membrane feeding with enriched gametocytes</u>

DMFA is conducted following an established protocol [18]. Venous blood is drawn in 10 mL heparin tubes from the study participants [19]. The blood is kept warm at 37°C until it is processed for enrichment (see below) or directly injected in water-jacketed glass membrane feeders that are connected to a heated water bath [17].

Magnetic column fractionation (magnetic activated cell sorting – MACS) or Percoll density gradient centrifugation is applied to enrich gametocytes out of venous blood prior to mosquito feeding, aiming to boost mosquito infection intensity [20]. Blood is kept warm throughout the process. The blood meal with enriched gametocytes is prepared and subsequently fed using water-jacketed glass membrane feeders.

Mosquitoes in cups and cages are allowed to ingest blood for 10 minutes through an artificial membrane feeder.

<u>Gametocyte enrichment procedures</u>

There are several described methodologies to enrich gametocytes out of whole blood [20]. We will use magnetic column fractionation and density gradient fractionation to determine the most effective way to enrich for *P. vivax* gametocytes and to maximize mosquito infection.

Magnetic column fractionation exploits the paramagnetic properties of mature gametocytes. Due to the accumulation of hemozoin (malaria pigment) mature gametocytes can be isolated out of whole

blood by passing through a column that is placed in a magnet. Following a wash with medium the column is removed from the magnet and gametocytes flow through. This technique is very efficient but not ideal for larger blood volumes (>10 mL). .
Density gradient fractionation uses the difference in density of infected and uninfected red blood cells to enrich for gametocytes. Whole blood is loaded on a density gradient made of Percoll, and centrifuged to allow collection of gametocytes that remain on top of the gradient.
With both methods gametocyte viability remains high. The Percoll concentration technique has been shown to achieve up to 40-fold enrichment in the number of *P. vivax* gametocytes that resulted in an increase in mosquito infection rates following membrane feeding from 10% to above 90%[6].

**Screening**

All potential participants who give informed consent and pass the study quiz will undergo a screening visit, which may take place within 28 days prior to inoculation of PvW1. The last possible screening date is three days before inoculation (Day I-3). In any case, all laboratory values need to be present and reviewed before inoculation. Information and consenting will be done before screening as described. When consent is obtained, the screening procedures indicated in the schedule of procedures (Appendix 1) will be undertaken. These procedures include β-HCG in serum in female participants, duffy antigen phenotyping, rhesus phenotyping, virology (HIV, HBsAg, WNV and HCV), complete blood count and biochemistry. For our last cohort, where we will generate mosquitoes that are planned to be used in clinical studies, we additionally take a detailed travel history and screen for additional mosquito borne diseases if indicated. If the appropriate blood test results for screening are available for a given participant due to screening for another study at the Radboudumc, these results may be used upon permission from the participant for assessing eligibility, provided the results date within the 28 days preceding enrolment in RaViCHMI1 (excluding tests where results would not change, such as genetic tests).
Abnormal clinical findings from the medical history, physical examination, or blood tests at any point in the study will be assessed using established local reference intervals. If a laboratory test is out of range, it may be repeated to ensure it is not a single occurrence. If an abnormal finding is deemed to be clinically significant, the participant will be informed, and appropriate medical care arranged with the permission of the participant. The decision to exclude the participant from enrolling in the study or to withdraw a participant from the study will be at the discretion of the Investigator. A 12-lead electrocardiogram is performed in each participant on screening. A detailed travel history will be obtained. Participants are only eligible if they pass all in- and exclusion criteria.

**Distribution of temperature and sleep measurement devices**

Once all test results from the screening visit are known and the participant is deemed eligible, a clinical visit will be organized where the participant will receive sleep-related questionnaires, self-monitoring, temperature and sleep measurement devices and a training on how to use it. Participants will start filling in the questionnaires and using the device on this day according to the instructions given by the investigators. Depending on timing, this visit may coincide with the Day-3 visit.

**Three days before inoculation (Day-3)**

Any new medical issues or symptoms that have arisen since screening will be assessed in case screening was before Day-3 (Day-3 is the latest possible screening date). Additional analyses include β-HCG in serum in female participants, complete blood count and biochemistry. Day-3 will be used as the baseline for subsequent analyses and samples for parasite quantitative polymerase chain reaction (qPCR), immunology, transcriptional and metabolic profiling will be taken. One serum aliquot (5 ml) is stored in -80°C until the end of the study to serve as a baseline sample if unexpected AEs, which require extended laboratory analyses, occur.
On Day-3, participants will be instructed on the use of the diary, home body temperature measurement as well as reporting of self-medication to the study team.

**Two days before inoculation (Day-2)**

No clinical visit. The participants start their diary. This continues until the end of the study. Latest on Day+49, self-monitoring equipment and data are collected.

**The day before inoculation (Day-1)**

In- and exclusion criteria will be reviewed, updates to the medical history will be recorded and all participants will be examined clinically. As for the other visits, sufficient time will be given to pose questions about the study, associated risks, malaria, treatment and all other matters that may have arisen.

**Day of inoculation (Day+0)**

An intravenous catheter will be placed and flushed with normal saline. For each participant, the inoculum must be injected within 4 hours of the inoculum being thawed, followed by a further normal saline flush. Before inoculation, baseline physical observations vital signs will be measured and recorded. Physical observations will be repeated at 15 minutes post-administration of the inoculum and again at 1 hour, in order to assess for immediate adverse reactions. A physician of the clinical team will be at the ward for the whole observation period. If the participant should show signs or symptoms of a transfusion reaction, the participant will be closely monitored for their vital signs, a blood culture will be taken and the intravenous line will be kept open in case symptoms need parenteral treatment. Oxygen and an emergency kit will be present at the unit, which is located within Radboudumc. Participants will be observed for at least 1 hour, however, this period may be extended if there are any clinical concerns. If there have been no symptoms or signs indicative of a transfusion reaction, the cannula will be removed after 1 hour.

Before leaving, participants will be informed about the (unlikely) possibility of a delayed transfusion reaction and will be provided with the 24-hour emergency mobile telephone number to enable them to contact a study physician in the event of concern. Additionally, a participation (medic-alert) ID card will be issued to each participant with information including antimalarial sensitivity of PvW1, study physician contact details and a request that the research team be contacted immediately in the event of illness/accident. If the participant does not have their own mobile telephone, they will be issued one for the duration of the study and counselled about the importance of keeping it switched on or checking the messages regularly. In addition, full contact details for each participant will be verified, including home address, home and work land-line telephone numbers where available and next-of-kin address, electronic mail address and telephone numbers. Participants must also provide the investigators with the name telephone number of at least one emergency contact.

**Day+1**

Participants will come to the site and will be seen by the clinical team to collect adverse events and a blood sample for immunological, transcriptional and metabolic measurements.

**Day+2 to Day+4 post inoculation**

Participants will be contacted daily by phone. In case they develop symptoms, they will be seen by a study physician and parasitaemia will be assessed by TBS and PCR. Additional diagnostic procedures may be performed as per discretion of the study team.

**Day+5 to Day+9 post inoculation**

Once daily parasitaemia measurements will be done by PCR. Frequency of PCR measurement may be increased if results are inconclusive, or a participant has symptoms. As part of the daily visit, vital signs will be measured, and adverse events (AEs) will be solicited. A TBS may be done, although parasitaemia is expected to be below the lower limit of detection (LLoD) until about Day+14. TBS results will be available within 12 hours. If related symptoms occur, TBS turn-over-time will be reduced to less than 6 hours. On Day+7 samples for immunology, transcriptional and metabolic profiling will be taken.

**Day+10 to Day+26 post inoculation**

Daily parasitaemia measurements by PCR and clinical visits continue as before. In addition, parasitaemia will be measured by TBS and the number of gametocytes will be assessed by PCR. Treatment with mepacrine (Group M) or piperaquine (Group P) will be initiated when a participant becomes TBS positive with >10 parasites/µL. For logistical reasons, in participants with no or easily tolerable symptoms, treatment will be started at the latest the following day. If a participant is TBS positive (at any parasitaemia) and has a body temperature >38.5°C and/or malaria-related symptoms that significantly interfere with daily life, treatment with mepacrine or piperaquine will be initiated immediately. Treatment decision is primarily based on TBS results with PCR results as backup. In case the TBS result is inconclusive, the PCR-measured parasite count (expressed in *P. vivax* genome copies/mL) will be used as replacement. For female participants a pregnancy test will be conducted prior to initiating gametocyte-sparing treatment. In case the female participant is tested positive during the pregnancy screen, curative treatment rather than gametocyte-sparing treatment will be initiated.

Group P will receive a single dose of 480 mg piperaquine, Group M participants are treated with 100 mg every 8 hours on the first day of treatment, followed by 100 mg daily for three days. On the day of treatment initiation, collection of blood samples for drug concentration measurement will start, following the sampling schedule detailed in Appendix 2. In addition, a sample for immunological analysis will be taken. During and after treatment daily monitoring of clinical symptoms and parasitaemia continues. On Day+24 or when parasitaemia is not controlled by treatment with mepacrine or piperaquine, participants will receive an end-of-study treatment (atovaquone-proguanil or artemether-lumefantrine). Treatment success will be assessed by PCR during the three days of treatment and the subsequent visit.

An additional blood sample for immunological, transcriptional and metabolomic measurements will be taken on Day+13 or the day of initiation of gametocyte-sparing treatment.

**Day+28**

Participants will be contacted by phone to ensure well-being of the participants after completion of end-of study treatment.

**Day+35 post inoculation**

Participants will be examined clinically, the absence of parasitaemia will be confirmed by PCR. Haematology and biochemistry values are obtained. A blood sample for immunological measurements will be taken. Diaries and equipment for temperature monitoring will be collected.

**Day+49 post inoculation**

The last scheduled visit of the study. Participants will be examined clinically, and haematology and biochemistry values are obtained. Questionnaires and equipment for sleep monitoring will be collected.

#### 8.4. Withdrawal of individual subjects

Study participants can leave the study at any time for any reason if they wish to do so without any consequences. The investigator can decide to withdraw a subject from the study for urgent medical reasons. Of note, volunteers who withdraw following inoculation of PvW1 but before end-of-study treatment will need a full course of antimalarial to clear the infection. This will be explained in detail during the informed consent process and is one of items in the study quiz that subjects have to pass before they are eligible.

#### 8.5. Replacement of individual subjects after withdrawal

Study participants withdrawing their informed consent before administration of a *P. vivax* blood inoculum can be replaced by back-up volunteers. Study participants withdrawing after having received a *P. vivax* blood inoculum will not be replaced.

#### 8.6. Follow-up of subjects withdrawn from treatment

Study participants that withdraw after having received a *P. vivax* blood inoculum but prior to being treated, require a full course of antimalarials. Treatment success will be monitored by PCR during the three days of treatment. A follow up visit will be proposed (and strongly suggested) to confirm parasite negativity.

#### 8.7. Premature termination of the study

The sponsor will terminate the study if there is evidence that continuation of the study will jeopardize the health or safety of the study participants. The sponsor will notify the accredited CCMO within 15 days of premature termination of the study including the reason for such an action. The investigator will take care that all subjects are kept informed premature termination of the study. All participants who have received the inoculum prior to premature termination of the study will be followed for safety until the full course of the final study treatment has been completed.

The study may be terminated prematurely by the sponsor and will take into account advice from:

- Local safety monitor
- Data safety monitoring board (DSMB)
- Clinical investigator
- Ethics committee

### 9. SAFETY REPORTING

#### 9.1. Temporary halt for reasons of subject safety

In accordance with section 10, subsection 4, of the Medical Research Involving Human Subjects Act (WMO), the sponsor will suspend the study if there is sufficient ground that continuation of the study will jeopardise the health or safety of the study participants. The sponsor will notify the accredited METC without undue delay of a temporary halt including the reason for such an action. The study will be suspended pending a further positive decision by the accredited METC. The investigator will take care that all study participants are kept informed.

The study may be discontinued by the sponsor and will take into account advice from:

- Local safety monitor
- Data safety monitoring board (DSMB)
- Clinical investigator
- Ethics committee

The study may be placed on hold in case:

- One or more participants experience a SAE that is at least possibly related to the inoculum or CHMI
- One or more participants experience a SAE that is at least possibly related to antimalarial treatment

#### 9.2. AEs, SAEs and SUSARs

##### 9.2.1. Adverse events (AEs)

Adverse events are defined as any undesirable experience occurring to a subject during the study, whether or not considered related to the study procedures. All adverse events reported spontaneously by the study participant or observed by the investigator or his staff will be recorded. Of note, a set of signs, symptoms and laboratory abnormalities (fatigue, malaise, headache, myalgia, arthralgia, back pain, nausea, anorexia, vomiting, diarrhoea, fever, feverishness, chills, rigor, sweats, tachycardia, hypotension, drowsiness, dizziness, lymphopenia and thrombocytopenia) are expected in a CHMI trial. They will be solicited and documented.

##### 9.2.2. Serious adverse events (SAEs)

A serious adverse event is any untoward medical occurrence or effect that

- results in death;
- is life threatening (at the time of the event);
- requires hospitalisation or prolongation of existing inpatients' hospitalisation;
- results in persistent or significant disability or incapacity;
- is a congenital anomaly or birth defect; or
- any other important medical event that did not result in any of the outcomes listed above due to medical or surgical intervention but could have been based upon appropriate judgement by the investigator.

An elective hospital admission will not be considered as a serious adverse event. If a participant feels unwell because of malaria symptoms and prefers to stay at the unit overnight for logistical reasons, the stay will not reported as a SAE. The same holds true for repeated sampling, drug intake or mosquito exposure in case they occur in the evening and the participant prefers to stay for logistical reasons (e.g. lack of transportation).
The investigator will report all SAEs to the sponsor without undue delay after obtaining knowledge of the events.
The sponsor will report the SAEs through the web portal *ToetsingOnline* to the accredited METC that approved the protocol, within 7 days of first knowledge for SAEs that result in death or are life threatening followed by a period of maximum of 8 days to complete the initial preliminary report. All other SAEs will be reported within a period of maximum 15 days after the sponsor has first knowledge of the SAE.

##### 9.2.3. Suspected unexpected serious adverse reactions (SUSARs)

Adverse reactions are all untoward and unintended responses to a product related to any dose administered.
Unexpected adverse reactions are SUSARs. The following conditions in this study meet the criteria of a SUSAR:

1. the event must be serious (see definition of SAE above);
2. there must be a certain degree of probability that the event is a harmful and an undesirable reaction to a non-investigational product administered in this study, regardless of the dose;

3. the adverse reaction must be unexpected, that is to say, the nature and severity of the adverse reaction are not in agreement with the product information.

The sponsor will report expedited the following SUSARs through the web portal *ToetsingOnline* to the METC:

- SUSARs that have arisen in this study which was assessed by the METC;
- SUSARs that have arisen in other clinical studies of the same sponsor and with the same product, and that could have consequences for the safety of the study participants involved in this study which was assessed by the METC.

The remaining SUSARs are recorded in an overview list (line-listing) that will be submitted once every half year to the METC. This line-listing provides an overview of all SUSARs from the involved non-investigational products, accompanied by a brief report highlighting the main points of concern.
The expedited reporting of SUSARs through the web portal *ToetsingOnline* is sufficient as notification to the accredited METC.
The sponsor will report expedited all SUSARs to the competent authorities in other Member States, according to the requirements of the Member States.
The expedited reporting will occur not later than 15 days after the sponsor has first knowledge of the adverse reactions. For fatal or life-threatening cases the term will be maximal 7 days for a preliminary report with another 8 days for completion of the report.

#### 9.3. Annual safety report

In addition to the expedited reporting of SUSARs and as part of the annual report, the sponsor will submit, once a year throughout the study, a safety report to the accredited METC.
This safety report consists of:

- a list of all suspected (unexpected or expected) serious adverse reactions, along with an aggregated summary table of all reported serious adverse reactions, ordered by organ system, per study;
- a report concerning the safety of the study participants, consisting of a complete safety analysis and an evaluation of the balance between the efficacy and the harmfulness of the non-investigational product.

#### 9.4. Follow-up of adverse events

All AEs will be followed until they have abated, or until a stable situation has been reached. Depending on the event, follow up may require additional tests or medical procedures as indicated, and/or referral to the general physician or a medical specialist.

SAEs need to be reported till end of study within the Netherlands, as defined in the protocol.

#### 9.5. Adverse event data collection

Signs and symptoms will be recorded in study diaries and reviewed during admission at the clinical facilities of the research centre, at all follow-up visits, and whenever a study participant reports signs or symptoms to the study physician between visits.

**Solicited adverse events**

Following and during administration of the inoculum, the following local and systemic AE will be solicited until Day+4:

- Local AE: pain, tenderness, erythema and induration and swelling at injection site.
- Systemic AE: headache, fatigue, fever, drowsiness, chills, myalgia, arthralgia, nausea, vomiting, diarrhoea.

From Day+5 onwards, signs and symptoms of systemic inflammation due to malaria will be recorded as solicited AEs. These include: fatigue, malaise, headache, myalgia, arthralgia, back pain, nausea, anorexia, vomiting, diarrhoea, fever, feverishness, chills, rigor, sweats, tachycardia, hypotension, drowsiness, dizziness, lymphopenia and thrombocytopenia.

**Unsolicited adverse events**

Unsolicited AE will be collected during the entire study period following the inoculum administration.

##### 9.5.1. Recording of adverse event data collection

For all adverse events, the study physician will record the diagnosis (i.e., disease or syndrome) rather than component signs, symptoms and laboratory values, if known. If the signs and symptoms are considered unrelated to an encountered syndrome or disease they should be recorded as individual AEs. If a primary AE is recorded, events occurring secondary to the primary event should be described in the narrative description of the case (e.g. primary AE = Orthostatic hypotension; secondary event may be fainting, head trauma, etc.). In case of hospitalizations for surgical or diagnostic procedures, the pre-existing condition should be recorded as the SAE, not the procedure itself. All adverse events/reactions (solicited and unsolicited), noted by the investigators will be accurately documented in the case report form by the investigators. For each event/reaction the following details will be recorded:

1. Description of the event(s)/reactions(s)
2. Date and time of occurrence
3. Duration
4. Intensity
5. Relationship with the intervention
6. Action taken, including treatment
7. Outcome

In addition, intensity of symptoms will be described as (1) mild, (2) moderate, or (3) severe according to the following scale:

- Mild (grade 1): awareness of symptoms that are easily tolerated and do not interfere with usual daily activity.
- Moderate (grade 2): discomfort that interferes with or limits usual daily activity.
- Severe (grade 3): disabling, with subsequent inability to perform usual daily activity, resulting in absence or required bed rest.

The investigator will attempt to establish a diagnosis of the event based on signs, symptoms and/or other clinical information. In such cases, the diagnosis should be documented as the AE/SAE and not the individual signs/symptoms. If an AE changes in intensity during the specified reporting period, a new description of the AE will be added. Interrupted AEs are registered as one AE if the interruption is <24 hours. When an AE/SAE occurs, it is the responsibility of the investigators to review all documentation (e.g. hospital progress notes, laboratory, and diagnostics reports) related to the event. The investigators will then record all relevant information regarding an AE/SAE in the corresponding case report form (CRF).

##### 9.5.2. Assessment of causality

The investigators are obliged to assess the relationship between study procedures and the occurrence of each AE/SAE. The investigators will use clinical judgement to determine the relationship. Alternative causes, such as natural history or the underlying diseases, concomitant therapy, other risk factors and the temporal relationship of the event will be considered and investigated. The relationship of the AE with the study procedures will be categorized as:

- Definitely: study procedures are the cause, another etiology causing the AE is not known.

- Probable: study procedures are the most likely cause: however, there are alternative reasonable explanations, even though less likely.
- Possible: there is a potential association between the event and applied study procedures, however, there is an alternative etiology that is more likely.
- Unlikely: a relationship to the study procedures is unlikely, however, it cannot be ruled out.
- Not related: a relationship to the study procedures cannot be reasonably established; another etiology is known to have caused the AE or is highly likely to have caused it.

When a regulatory authority requests a binary classification (related vs. unrelated), definitely, probably and possibly related are considered to be “related”, while not related and unlikely related are considered to be “unrelated”. Thus, a study-related AE refers to an AE for which there is a possible, probable or definite relationship to the study procedures. The investigator will use clinical judgement to determine the relationship.

#### 9.6. Data Safety Monitoring Board (DSMB)

An independent DSMB composed of three independent individuals will be appointed. The DSMB will include a local safety monitor, and two experts nominated by the investigators. The DSMB will be established for the purpose of periodically reviewing and evaluating accumulated data on the safety of study participants, parasitaemia, gametocytaemia, and the study conduct and progress in general. The DSMB shall provide independent, non-binding advice on safety and ethics to the investigators and sponsor. The responsibilities and procedures of the DSMB members are defined in the DSMB Charter.

The advice(s) of the DSMB will only be sent to the sponsor of the study. Should the sponsor decide not to fully implement the advice of the DSMB, the sponsor will send the advice to the reviewing METC, including a note to substantiate why (part of) the advice of the DSMB will not be followed.

**Local safety monitor**

For this study, a local safety monitor (LSM) will be appointed, who will be involved in the review of severe and serious AE and study participants’ safety. The LSM is independent of the sponsor and the investigator. The LSM is notified of all SAEs, as well as at least all grade 3 AE probably or definitely related to the study procedures and persisting at grade 3 for >48 hours.

**Safety meetings**

The DSMB will review safety data upon completion of Group P and Group M, prior to initiating the next cohort. The chair of the DSMB will determine whether a (digital) meeting will be held, or whether a recommendation from all members may be formalized through e-mail. In addition, safety data for all participants will be assessed by the DSMB at the end of the study. An ad-hoc DSMB meeting may be convened at any time or at the request of the DSMB chair, investigators or local safety monitors to review safety data from study participants who meet any of the holding rules as specified in the protocol, or if otherwise deemed necessary.

**Safety reports**

A safety report will be prepared by the clinical investigators for review by the board prior to each scheduled data review. These reports will provide at a minimum the following information:

- Actual data and subject status data with regard to completion of/discontinuation from the study.
- Summaries of solicited AEs, classified by severity.
- Unsolicited AEs (including SAEs), categorized by Medical Dictionary for Regulatory Activities (MedDRA) coding, severity and relatedness to study vaccine.
- Safety laboratory test results outside of normal institution reference ranges and considered clinically significant, classified by severity grading scale (irrespective of whether assessed as AEs).
- Any new or updated AEs that have met the holding rules.

The DSMB will review the safety data within 5 working days. The DSMB will summarize their recommendations to the study sponsor as to whether there are safety concerns and whether the study should continue without change, be modified, or be terminated. If at any time a decision is made to permanently discontinue the study in all study participants, the sponsor will notify the accredited METC expeditiously.

**Holding rules**

If any of the participants meet a holding rule (see below), initiation of further cohorts will depend on a positive safety data review by the DSMB. The following holding rules apply:

- One or more participants experience a SAE that is at least possibly related to the inoculum or CHMI

- One or more participants experience a SAE that is at least possibly related to antimalarial treatment

The study site member first aware of the event meeting the holding rule will notify the Principal Investigator (PI) and the LSM of the corresponding site. The PI will alert the appropriate parties including the sponsor. The DSMB will be notified within 24 hours. An ad-hoc DSMB review will be performed. The following considerations must be discussed:

- Relationship of the AE or SAE to the study procedure
- If appropriate, additional screening or laboratory testing is provided to other study participants to identify participants who may develop similar symptoms
- If any study related SAE is not listed on the current informed consent form (ICF), the PIs will revise the ICF and upon approval of the updated ICF by the accredited METC, study participants will be asked to provide consent on the new ICF.

Initiation of the next cohort may resume only if the local safety monitor, PI, DSMB and the sponsor agree it is safe to resume the study. Temporary halts for reasons of subject safety need to be reported immediately to the accredited METC. Restart of the study activities following a temporary halt for reasons of subject safety requires a positive decision from the accredited METC.

The PI, sponsor or regulators may stop or suspend the initiation of new study cohorts at any time. All participants who have received the inoculum already will be followed for safety until the full course of the final study treatment has been completed.

### 10. STATISTICAL ANALYSIS

RaViCHMI1 is an exploratory study that is intended to optimize an existing human infection model for the use in transmission research and will allow the assessment interventions intended to inhibit transmission, such as transmission-blocking vaccines and drugs. The primary objective is to reproducibly infect mosquitoes with *P. vivax* during CHMI. To achieve this, parasitaemia that is associated with significant symptoms would be required [9]. To improve tolerability of CHMI while keeping transmission to mosquitoes high, participants will be treated early during infection with gametocyte-sparing antimalarials. In addition, it will be assessed if gametocyte concentration techniques can be used to further increase transmission. The study is not intended as confirmatory but to inform subsequent interventional studies.

Descriptive statistics on infection rate of participants and mosquitoes as well as tolerability and safety data will be reported. In addition, the kinetics of parasites during CHMI as well as drug concentrations, metabolic and behavioural parameters will be listed.

#### 10.1. Primary study parameter(s)

The proportion of infected mosquito infections will be estimated from approximately 30-40 mosquitoes (only fed mosquitoes will be dissected) at each respective feeding time point using three different methods (DFA, DMFA and DMFA following gametocyte enrichment). DMFAs will be done at all time points and DFA 3-4 times during peak gametocytaemia. For primary analysis a mixed effects logistic regression model with participant as random effect will be used.

#### 10.2. Secondary study parameter(s)

Safety and tolerability will be tabulated and graphically depicted using a set of solicited symptoms/AEs and MedDRA-coded unsolicited AEs.

#### 10.3. Other study parameters

The difference between the three mosquito feeding methods (DFA, DMFA and DMFA following gametocyte enrichment) and the effects of treatment and timing on infectivity will be analysed by a generalized additive mixed model. Explorative endpoints will be presented as descriptive statistics and using best practices of the different disciplines (molecular, cytometry, transcriptomic, proteomic and epigenetics data, measurement of sleep and metabolism using wearables and questionnaires). Drug concentration over time and its effect on infection kinetics, gametocytaemia and gametocyte sex-ration will be modelled using generalized mixed models.

##### 10.4. Interim analysis (if applicable)

No formal interim analysis will be done. Data from Group P and Group M will be used to inform Group O.

### 11. ETHICAL CONSIDERATIONS

#### 11.1. Regulation statement

This study will be conducted in accordance with the ethical principles from the latest Fortaleza revision of the Declaration of Helsinki (2013). Further, this study is conducted in compliance with the WMO, the International Council for Harmonisation of Technical Requirements for Pharmaceuticals for Human Use (ICH) Good Clinical Practice (GCP), and local regulatory requirements. The investigators are responsible for obtaining ethical approval of the protocol in compliance with local law before the start of the study. Any subsequent amendments also require ethical approval prior to implementation.

#### 11.2. Recruitment and consent

Upon approval of the study by the responsible ethics committee, healthy adults will be recruited to participate in the study. Advertisements will be placed in prominent areas on university campuses and other public places as well as on the intranet of the university and on dedicated social media platforms. The advertisement will indicate a telephone number to call and an e-mail address to contact for requesting further information. The webpage of the investigator site will provide an electronic registration form. When seemingly suitable subjects contact investigators via e-mail, telephone or the online registration form, they will be invited to an information meeting (in-person or digital) during which the study will be explained to them by a study physician. Directly after the meeting they will be provided with documents to review at home (the participant information leaflet, the informed consent form, the application form and the insurance text). During and after the meeting there will be time for questions. The possibility for follow-up discussions will be provided if requested by a volunteer. Volunteers are given sufficient time to consider participation. Healthy adults who are still interested in participating after attending the information session and considering their participation will be asked to fill in the application form and will be invited to come for a screening visit. Seemingly eligible volunteers shall only be included in the study after providing written informed consent on the informed consent form. Informed consent must be obtained by a study physician and before conducting any study-specific procedures (i.e. all of the procedures described in the protocol, including screening procedures). The process of obtaining informed consent shall be documented in the study participant's source documents. During the screening visit, the volunteer will complete a quiz for verification that the provided study information is understood. Additionally, a medical questionnaire will be completed and answers will be discussed. Inclusion and exclusion criteria will be checked. Again, the study physician will answer any questions the volunteer has. The voluntary nature of study participation and the possibility of withdrawal from the study, at any time, without penalty and without any declaration of the reason will be pointed out to the volunteer. The requirement for a full course of antimalarials for withdrawal after CHMI will be discussed. The investigators will be responsible for providing adequate verbal and written information regarding the objectives and procedures (including self-monitoring tools) of the study, the potential risks involved and the obligations of the study participants. The potential study participants will be informed that they will not gain health benefits from this study. Informed consent will be dated and signed by the study physician performing the interview, and by the study participant. The study participant will be provided with a signed copy of the document by which informed consent has been given. Trainees or other students who might be dependent on the investigators or the study group will not be eligible for study participation.

##### 11.3. Objections by minors or incapacitated subjects

Not applicable.

##### 11.4. Benefits and risks assessment, group relatedness

The need for developing tools to test vaccines against *P. vivax*, as requested in the World Health Organization (WHO) malaria technology roadmap, to protect populations at risk requires to be balanced with the potential risks and discomforts for the study participants. Risks of the study can be divided into risks associated with the inoculum, *P. vivax* malaria, antimalarial treatment, mosquito bites, and transmission of malaria. There are no direct benefits to participation in this study for the study participants. The societal benefit of the study is to develop a tool that will accelerate efficacy testing on drug and vaccine interventions against *P. vivax*.

All partners in this proposal are aware of and follow the relevant national and international rules and regulations as they pertain to access to data and material of human origin and clinical research. International agreements such as the Declaration of Helsinki will be observed and respected.

##### 11.5. Compensation for injury

The sponsor/investigator has a liability insurance which is in accordance with article 7 of the WMO. The sponsor (also) has an insurance which is in accordance with the legal requirements in the Netherlands (Article 7 WMO). This insurance provides cover for damage to research subjects through injury or death caused by the study.

The insurance applies to the damage that becomes apparent during the study or within 4 years after the end of the study.

The insurance offers a maximum coverage of:

1. € 650.000,-- (i.e. six hundred and fifty thousand euro) for death or injury for each subject who participates in the research;
2. € 5.000.000,-- (i.e. five million euro) for death or injury for all subjects who participate in the research;
3. € 7.500.000,-- (i.e. seven million five hundred thousand euro) for the total damage incurred by the organisation for all damage disclosed by scientific research for the sponsor in the meaning of said Act in each year of insurance coverage.

##### 11.6. Incentives

Enrolled subjects will receive up to 1400 Euro in compensation for their time and for the inconveniences of taking part in this study. These amounts are based on predefined criteria following a Standard Operating Procedure:

| | |
|---|---|
| • Screening and inclusion | 125 Euro |
| • Controlled Human Malaria Infection | 125 Euro |
| • In-person follow up visits | 25 Euro |
| • Mosquito feeding visits (and other prolonged visits) | 50 Euro |

Travel expenses for study participants living outside of Nijmegen will be additionally reimbursed. Compensation will not be provided to volunteers who are not enrolled (i.e. screen failures). Eligible volunteers who are enrolled at the inclusion visit as back-ups, will be compensated 225,- Euro for each inclusion visit. If a study participant withdraws from the study prior to study completion, reimbursement proportional to the number of visits attended will be provided. These compensation amounts are reasonable and in line with Dutch common practice. In case of unexpected medical complications, there will be access to state-of-the-art medical treatment with full costs covered by the insurance of Radboudumc.

##### 11.7. Other ethical considerations

###### 11.7.1. Incidental findings

Although the probability of incidental findings in the proposed study is minimal due to the applied methodologies, the investigators will, in case of incidental findings, discuss these findings in the *Commissie Nevenbevindingen* of Radboudumc, a committee specialised in dealing with incidental findings. The committee will clarify the significance of the incidental finding and advise the study team whether the finding should be shared with the participant.

###### 11.7.2. Environment

All work performed during this project will be performed using the highest standards of environmental protection. This includes a safe and environmentally friendly manner, i.e. all waste will be treated following national legislation.

### 12. ADMINISTRATIVE ASPECTS, MONITORING AND PUBLICATION

The study protocol, informed consent form, study participant information leaflet, study advertisement, product information and any other documents required will be submitted for ethical approval before the research study is started.

#### 12.1. Handling and storage of data and documents

Data management activities from project set-up through data lock and transfer will follow a study-specific data management plan (DMP). Designated and trained study staff will enter the data required by the protocol into electronic case report forms (eCRF). All data will be recorded, handled and stored in a way that allows its accurate reporting, interpretation and verification. The study monitor will review the data entered into the eCRFs by investigational staff for completeness and accuracy and will instruct the site personnel to make any required corrections or additions. Queries are made during each monitoring visit and investigators are required to respond to the queries and confirm or correct the data. Medical history and current medical conditions will be coded using the ICD-10 terminology. AEs will be MedDRA-coded.

##### 12.1.1. Source Data

Only authorized study staff and representatives of the sponsor and responsible ethics committee agencies may have direct access to source documents containing study data. All information in original records and certified copies of original records, clinical findings, or observations will be considered source data for this study. Source data for this study will include biographical, medical history, clinical (signs, symptoms, prescribed and non-prescribed medical treatments), safety and laboratory data, and memory aids produced by the study participants. Paper-based source documents will be filed in the investigator’s clinical file. Since all study participants will be healthy, there is no medical file at study start. Medical files will be added in case of medical consultations or hospitalizations during the study conduct. In this case the medical file will also be considered source data.

All data collected by the investigators is reported in eCRF. As with all parts of the eCRF, there is an audit trail in place to register every data entry. The investigator will also keep the original informed consent form signed by the study participant (in addition a signed copy is given to the study participant). The investigator’s team will store all data collected in secure and access limited areas and treat these data as confidential material. Documents and data pertaining to the study will be kept in a locked cabinet under the responsibility of the investigator. Periodic monitoring visits will ensure that the data is safe and stored in a secure place and that only authorized study staff have access to the data. Source data from this study will be stored for 25 years, in line with the *Nederlandse Federatie van Universitair Medische Centra* (NFU) guideline. Biological samples from this study will be stored for a maximum of 15 years for research related to this study. New immunological or molecular tests may become available in the future that could strengthen or validate this research or help find important new findings. Should material be used for research not related to RaViCHMI1, prior permission from the ethics committee must be sought.

###### 12.1.2. Data protection & safeguarding:

In this study personal data from the study participants will be collected. All parties involved and in particular the clinical research sites will adhere to the European Union (EU) Regulation 2016/679: General Data Protection Regulation (GDPR). All parties further agree to adhere to the principles of medical confidentiality in relation to clinical study participants and shall not disclose the identity of study participants to third parties without prior written consent of the participant.

Further, the DMP will be reviewed by a Data Protection Officer (DPO). Implementation of the DMP requires prior approval from the DPO. The DPO will ensure all necessary data protection measures are implemented, data access and storage is in line with standardized data security measures and adherence to EU GDPR is guaranteed. Contact details of the DPO will be shared with the study participants during the informed consent process.

Researchers will take every reasonable step to protect the confidentiality of the study participants' health and personal information and to prevent misuse of this information. Upon inclusion into the study, each study participant will be assigned a unique identification code (pseudonymization) that will be used for the data collection on the clinical record forms, for data entry, data transfer, and data analysis.

Participants' medical records, Informed Consent documents, and all documents displaying personal identifiers will be kept in a locked cabinet at the investigator's site. Only selected study team members will have access to these documents containing personal identifiers. Access rights are documented on the delegation of responsibilities log. The identification key linking the unique identifier codes with the participant's direct identifiers (e.g. name of the study participant) will be stored in an access-limited and access-controlled folder and separate from any other study data. Again, only selected study team members will have access to the identification key log.

Researchers will also make sure biological samples are handled with care and that study participants' privacy is protected during collection, transfer, processing, and storage. The collected biological samples will be pseudonymized and labelled with the subject's unique identification code before being sent to a laboratory for analysis.

Data management procedures will protect the confidentiality of all individual data and ensure compliance with Dutch and European regulations: names will not be listed nor any set of demographic data, which could allow specific identification of study participants (e.g. only year of birth but not date of birth, etc.).

Disclosure of personal information from the study to third parties may only occur for purposes of study monitoring, audits and inspections. Study participant's confidentiality and welfare will always be maintained as the highest priority. Study participants will be informed about all parties having access to their personal information during the informed consent process.

From a technical point of view, data protection will be ensured by the Sponsor that will be in charge of providing tools for safeguarding the identity of study participants and protecting the data and study databases, following the standardized data security procedures. This includes:

- Physical Safety: Access to computer servers is restricted to authorized and trained personnel.
- Protection of computer servers: Access to the room hosting the database server is controlled. Only system team staff is allowed to enter this room and entries are logged. Computer servers are protected against power failure by UPS devices. Computer servers are kept at a constant temperature and humidity environment, and are monitored by fire detectors. Remote access to computer servers is strictly controlled by a high security level password provided to authorized personnel only. Servers are protected from outside access by a wired firewall.
- Backup procedures: Each server is backed-up daily with a weekly/monthly additional backup. Daily back-ups are kept in a fire-resistant storage place. A weekly back-up is stored in another building remotely from the server computer room. A monthly back-up is also kept.
- Authentication: Access to the study database will be controlled by user-specific passwords.
- Server connection protocols: Privileged accounts (root, admin or system for example) will be restricted to study database system developers.

Sending data outside of the European Union

During this study we may send pseudonymized data to the United Kingdom (UK) and United States (US). Data will be shared under EU Standard Contractual Clauses (EU SCCs). The UK endorses the EU SSCs under their own national data protection legislation. Data transfer to the US will also be done under the EU SCCs.

#### 12.2. Monitoring and Quality Assurance

##### 12.2.1. General considerations

Before study initiation, the study protocol and eCRFs together with relevant standard operating procedures (SOPs) will be reviewed by the sponsor, the investigators and their staff. The study monitor will visit the site prior to study initiation, during and after completion of the study to check the completeness of records, the accuracy of entries on the eCRFs, the adherence to the protocol and to Good Clinical Practice and the progress of enrolment.

The investigator will maintain source documents for each participant in the study, consisting of case and visit notes containing demographic and medical information, laboratory data, study participant's diaries, and the results of any other tests or assessments. All information on eCRFs must be traceable to these source documents in the study participant's file. The electronic data capture (EDC) maintains an audit trail.

The investigator will give the study monitor access to all relevant source documents to confirm their consistency with the eCRF entries. This study is classified as high risk. Therefore, the monitor will perform full verification for the presence of informed consent, adherence to the inclusion/exclusion criteria and documentation of SAEs. The recording of data that will be used for all primary and safety endpoints will be assessed for 100% of included study participants.

##### 12.2.2. Study monitoring

To ensure that the study is conducted in accordance with the Declaration of Helsinki and ICH-GCP and applicable regulatory requirements, monitoring responsibilities will be provided by independent and experienced study monitors. A site initiation visit will be conducted prior to beginning of the study. Interim monitoring visits will be conducted as laid out in the monitoring plan. Further, a final closeout monitoring visit will be performed. During the course of the study, monitors will verify 1) compliance with the study protocol, 2) completeness, accuracy, and consistency of the data, 3) non-investigational product storage and accountability and 4) adherence to the Declaration of Helsinki, ICH-GCP and applicable regulations. As needed and when appropriate, the study monitors will also provide clarifications and additional training to help the site resolve issues identified during the monitoring visit. As appropriate and informed by risk assessment, remote centralized monitoring activities may be considered in lieu of or to supplement onsite monitoring. These may include analysis of data quality (e.g. missing or inconsistent data), identification of data trends not easily detected by onsite monitoring, and performance metrics (e.g. screening or withdrawal rates, eligibility violations, timeliness, and accuracy of data submission).

The extent and frequencies of the monitoring visits will be described in a monitoring plan. The investigator will be notified in advance of scheduled monitoring visits. The monitor should have access to all study related facilities, study participant's records, non-investigational product accountability, and other study-related records needed to conduct monitoring activities. The study monitor will share the findings of the monitoring visit, including any corrective actions, with the investigator. The investigator and the monitors must cooperate to ensure that any problems detected in the course of these monitoring visits are resolved in a predefined time frame. All queries must be resolved prior to database lock. Essential documents must be filed in the site study file on an ongoing basis and be available for review by the study monitor.

##### 12.2.3. Independent Auditing

Sponsor representatives may audit the study to ensure that study procedures and data collected comply with the protocol and applicable SOPs at the clinical site and that data are correct and complete. The investigators will permit auditors to verify source data validation of the regularly monitored clinical study. The auditors will compare entries in the eCRFs with the source data and evaluate the study site's adherence to the clinical study protocol, ICH GCP guidelines, and applicable regulatory requirements.

##### 12.2.4. Inspections

The site PI must be aware that regulatory authorities may wish to inspect the site records to verify the validity and integrity of the study data and protection of human research participants. The site PI must make the relevant records available for inspection and will be available to respond to reasonable requests and audit queries made by authorized representatives of regulatory agencies.

##### 12.3. Amendments

A 'substantial amendment' is defined as an amendment to the terms of the accredited METC application, or to the protocol or any other supporting documentation, that is likely to affect to a significant degree:

- the safety or physical or mental integrity of the study participants;
- the scientific value of the study;
- the conduct or management of the study; or
- the quality of the data or safety of the CHMI.

All substantial amendments will be notified to the accredited METC.
Non-substantial amendments (e.g. typing errors and administrative changes like changes in names, telephone numbers and other contact details of involved persons mentioned in the submitted study documentation) will not be notified to the accredited METC but will be recorded and filed by the sponsor.

##### 12.4. Annual progress report

The sponsor/investigator will submit a summary of the progress of the study to the accredited METC once a year. Information will be provided on the date of inclusion of the first subject, numbers of subjects included and numbers of study participants that have completed the study, serious adverse events/ serious adverse reactions, other problems, and amendments.

##### 12.5. Temporary halt and (prematurely) end of study report

The investigator/sponsor will notify the accredited METC of the end of the study within a period of 8 weeks. The end of the study is defined as the last participant's last visit.
The sponsor will notify the accredited METC immediately of a temporary halt of the study, including the reason for such an action.
In case the study ends prematurely, the sponsor will notify the accredited METC within 15 days, including the reasons for the premature termination.
Within one year after the end of the study, the investigator/sponsor will submit a final study report with the results of the study, including any publications/abstracts of the study, to the accredited METC.

##### 12.6. Public disclosure and publication policy

The study will be entered into a primary registry of the International Clinical Trials Registry Platform (ICTRP) network of the WHO. The final report will be prepared by the investigators. It will be signed by the sponsor and the principal investigator. The investigators will publish the results of the study, preferably in a peer-reviewed journal.

#### Appendix 1: Schedule of procedures

| | Screening | CHMI-3 | CHMI-2 | CHMI-1 | CHMI | CHMI +1 | CHMI +2 to +4 | CHMI +5 to +9 | CHMI +10 to +26 | CHMI +28 | CHMI +35 | CHMI +49 |
|---|---|---|---|---|---|---|---|---|---|---|---|---|
| Days [in relation to day of inoculation] | -28 to -3 | -3 | -2 | -1 | 0 | +1 | +2 to +4 | +5 to +9 | +10 to +26 | +28 | +35 | +49 |
| **Clinical Procedures** | | | | | | | | | | | | |
| Clinic (C) or phone (P) visit or home data collection (H) | C | C | H | C | C | C | P | C | C | P | C | C |
| Written informed consent | X | | | | | | | | | | | |
| Study Quiz | X | | | | | | | | | | | |
| In-/exclusion criteria | X | | | X | | | | | | | | |
| Demographic & medical history | X | X | | X | | | | | | | | |
| Full physical exam | X | | | X | | | | | | | X | X |
| Vital signs | X | X | | X | X | | | X | X | | X | X |
| ECG | X | | | | | | | | | | | |
| Travel history | X | | | | | | | | | | | |
| Concomitant medication | X | X | | | X | | | X | X | | X | X |
| Review contraindications | | | | | X | | | | | | | |
| Address & next of kin | X | | | | X | | | | | | | |
| Clinical assessment (and/or collection of AEs) | | X | | X | X | X | | X | X | | | |
| Diary | | X[1] | X | X | X | X | X | X | X | X | X[3] | X[3] |
| Sleep questionnaires | | X[1] | X | X | X | X | X | X | X | | | |
| Home body temperature | | X[1] | X | X | X | X | X | X | X | | X[3] | X[3] |
| Sleep monitoring | | X[1] | X | X | X | X | X | X | X | | X[3] | X[3] |
| ID card | | | | | X[1] | | | | | | | |
| Inoculation | | | | | X | | | | | | | |

| | Screening | CHMI-3 | CHMI-2 | CHMI-1 | CHMI | CHMI +1 | CHMI +2 to +4 | CHMI +5 to +9 | CHMI +10 to +26 | CHMI +28 | CHMI +35 | CHMI +49 |
|---|---|---|---|---|---|---|---|---|---|---|---|---|
| Gametocyte-sparing treatment | | | | | | | | X[4] | X[4] | | | |
| Curative treatment | | | | | | | | | X[5] | | | |
| **Clinical sampling** | | | | | | | | | | | | |
| Drug screening | X | | | | | | | | | | | |
| Duffy & rhesus antigen screening | X | | | | | | | | | | | |
| Pregnancy test | X | X | | | | | | | X[6] | | | |
| Virology (HIV, HBV, HCV,WNV) | X | | | | | | | | | | | |
| Hematology | X | X | | | | | | | X[7] | | X | X |
| Biochemistry | X | X | | | | | | | X[7] | | X | X |
| qPCR | | X | | | | | | X | X | | X | |
| Thick blood smear | | | | | | | | X[2] | X | | | |
| **Research sampling** | | | | | | | | | | | | |
| Baseline serum aliquot | | X | | | | | | | | | | |
| Immunology/Transcriptomics/Metabolites | | X | | | | X | | X[8] | X[8] | | X | |
| Serum drug levels | | | | | | | | | X[9] | | | |
| **Entomology sampling** | | | | | | | | | | | | |
| Feeding assays | | | | | | | | | X[9] | | | |
| **Cumulative amount of blood** | | | | | | | | | **<500 ml** | | | |

1…hand out to study participant and provide instructions on usage; 2…optional; 3…collection from study participant; 4…Initiation of treatment depends on meeting treatment criteria as defined in the study protocol; 5…initiation on CHMI+24 or as defined in the study protocol; 6…on day of initiating gametocyte-sparing treatment;7…on CHMI+10 and day of initiation of gametocyte-sparing treatment; 8...on two time points during follow-up (CHMI+7 and CHMI+13 or the day of initiation of gametocyte-sparing treatment); 9…see Appendix 2 for further details

#### Appendix 2: Sampling schedule for measurements depending on initiation of gametocyte-sparing treatment

| Assay | Sampling timepoints | Comments |
|---|---|---|
| Entomology timepoints for the first group – these are subceptible to adaptation based on the results. | | |
| Direct Skin Feeding (option 1) | GST -0 hours | On the starting day of GST the participants will have two DFAs, if possible. One in the morning and one in the evening. |
| | GST -0 hours | |
| | GST +2 days | |
| Direct Skin Feeding (option 2) | CHMI +13 days | Option 2 if no GST is required |
| | CUT -0 hours | |
| DMFA and DMFA enriched | Day+11 | |
| | Day+13 | Daily DMFA until parasitemia <100 parasites/mL |
| | GST | In case that treatment starts before Day+13, daily DMFA will start on the day of GST |
| Serum drug levels | | |
| Mepacrine treatment | GST -0 hours | |
| | GST + 1 day | |
| | GST + 2 days | |
| | GST + 3 days | |
| | GST + 7 days | |
| | Day+26 | |
| | Day+35 | |
| | Day+49 | |
| Piperaquine treatment | GST -0 hours | |
| | GST +3 hours | |
| | GST + 6 hours | Might be delayed by up to 5 hours |
| | GST + 8 hours | Might be delayed by up to 5 hours |
| | GST +24 hours | |
| | GST + 7 days | |
| | Day+26 | |
| | Day+35 | |

GST…first dose of gametocyte-sparing treatment; CUT…first dose of curative treatment
