## Supplementary appendix for "A *Plasmodium vivax* controlled human infection and transmission model to evaluate interventions across the life cycle"

**Figure S1.** Microscopy images of thick blood smears.

1A. Multiple young ring-stages visible within the same field.

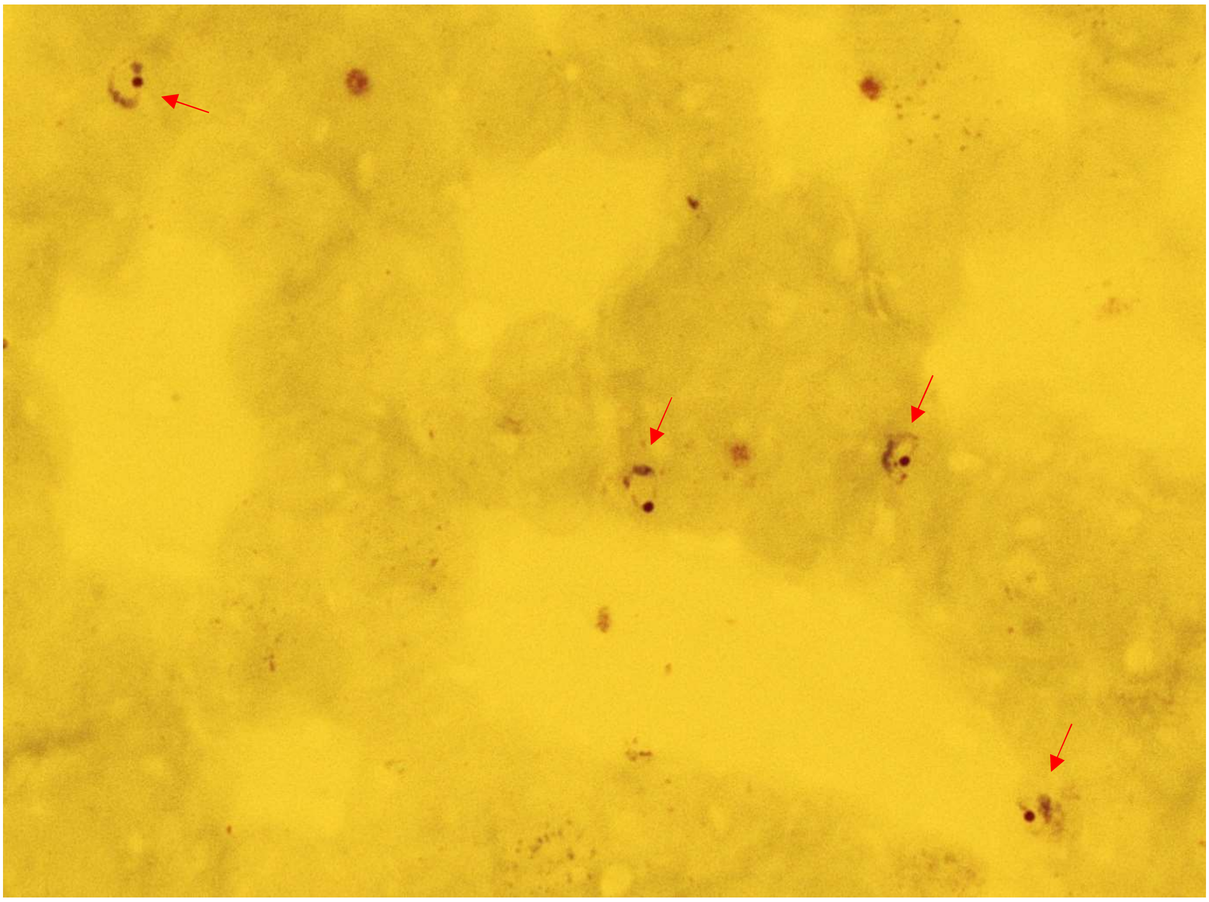

1B. Multiple older stages visible within the same field: schizont (green), gametocyte (red) and late stage trophozoite (blue).

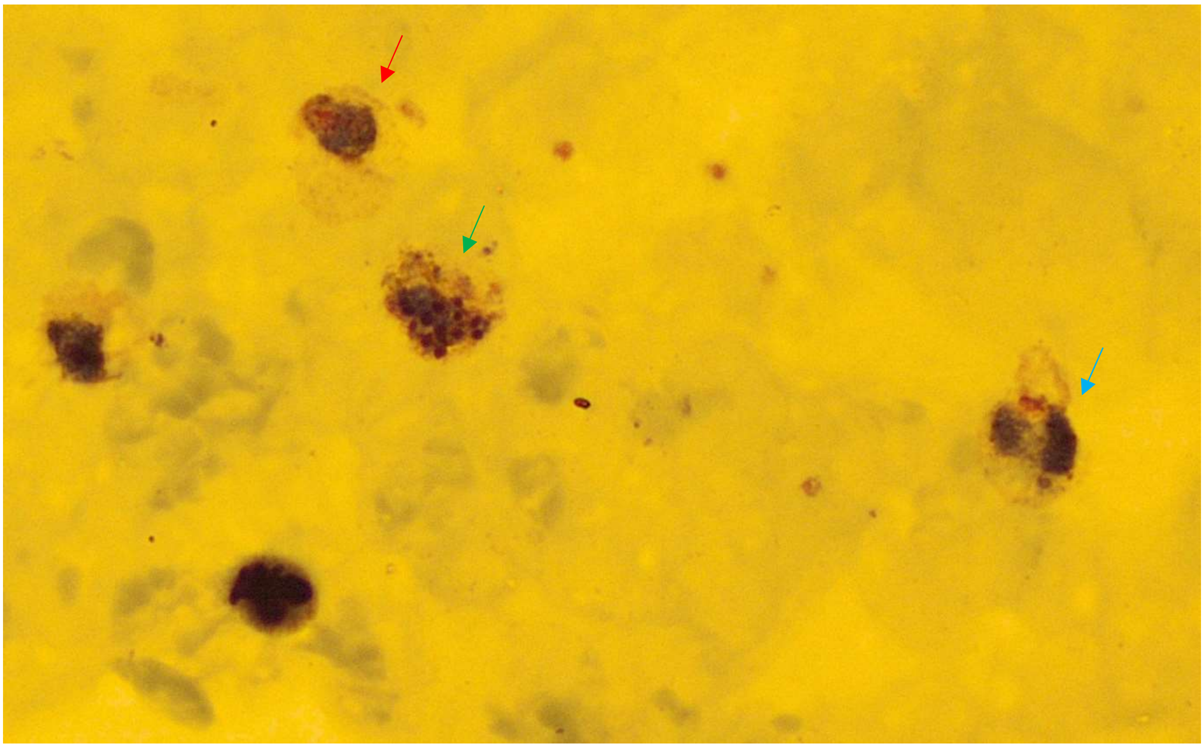

**Figure S2.** *P. vivax* sporozoites per oocyst over time following infectious feeding.
Sporozoite output was normalized to the mean oocyst count per mosquito to account for differences in midgut infection intensity across feeding experiments. Each point represents an individual mosquito cage on a given day post infectious blood meal. Violin plots show the distribution of sporozoites per oocyst for days with ≥3 observations (13–15 dpi). White diamonds indicate the mean per day, and the connecting line illustrates descriptive temporal trends.

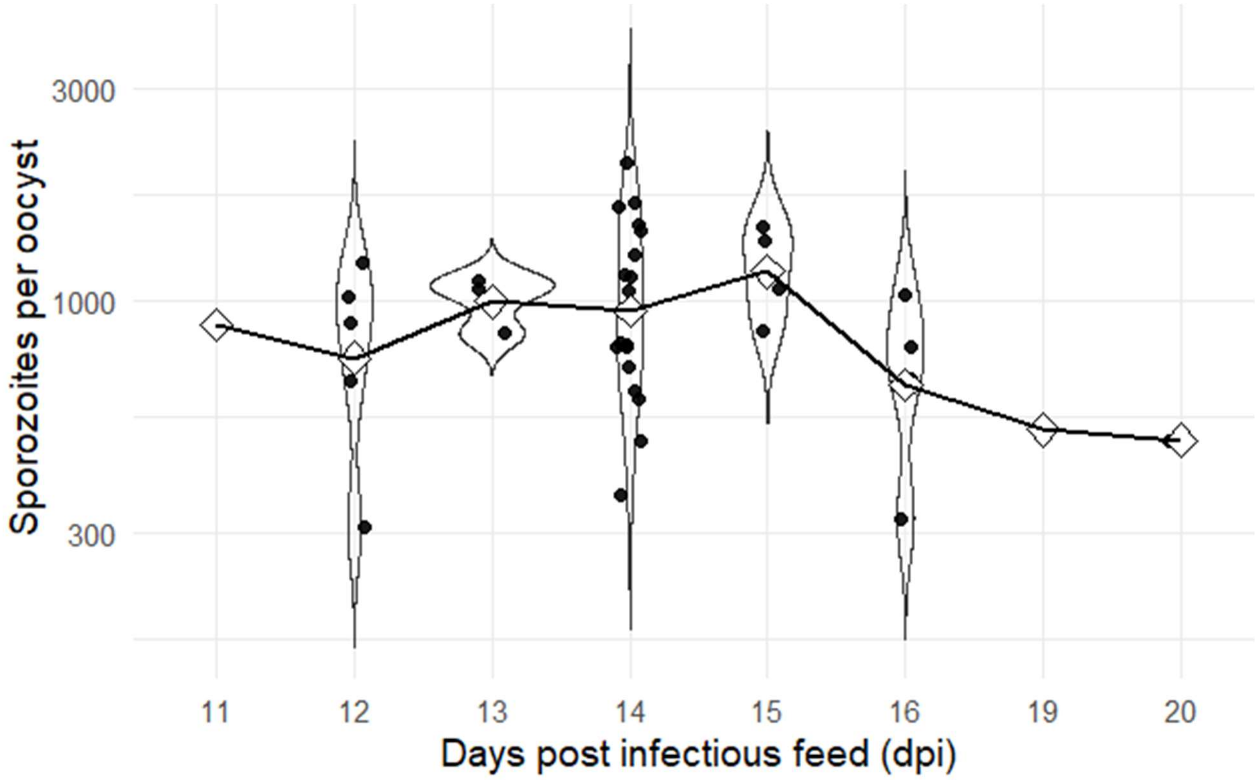

**Table S1.** Overview of solicited AEs after atovaquone-proguanil initiation.

| Day | Symptom | Grade | Count | Participants (n/N) |
|---|---|---|---|---|
| T+1 | Loss of appetite | 1 | 1 | 1/12 |
| T+1 | Loss of appetite | 2 | 1 | 1/12 |
| T+1 | Fatigue | 1 | 1 | 1/12 |
| T+1 | Fatigue | 2 | 2 | 2/12 |
| T+1 | Fever | 1 | 1 | 1/12 |
| T+1 | Headache | 1 | 3 | 3/12 |
| T+1 | Malaise | 1 | 2 | 2/12 |
| T+1 | Malaise | 2 | 2 | 2/12 |
| T+1 | Nausea | 1 | 1 | 1/12 |
| T+1 | Sweats | 1 | 1 | 1/12 |
| T+1 | Vomiting | 1 | 1 | 1/12 |
| T+1 | Vomiting | 2 | 1 | 1/12 |
| T+2 | Diarrhoea | 1 | 1 | 1/12 |
| T+2 | Dizziness | 1 | 1 | 1/12 |
| T+2 | Drowsiness | 1 | 1 | 1/12 |
| T+2 | Fatigue | 1 | 2 | 2/12 |
| T+2 | Headache | 1 | 2 | 2/12 |
| T+2 | Muscle ache | 1 | 1 | 1/12 |
| T+2 | Nausea | 1 | 2 | 2/12 |
| T+3 | Loss of appetite | 1 | 1 | 1/12 |
| T+3 | Sweats | 1 | 1 | 1/12 |
| T+4 | Drowsiness | 1 | 1 | 1/12 |

**Figure S3.** Image of acute allergic urticaria taken minutes after direct skin feeding

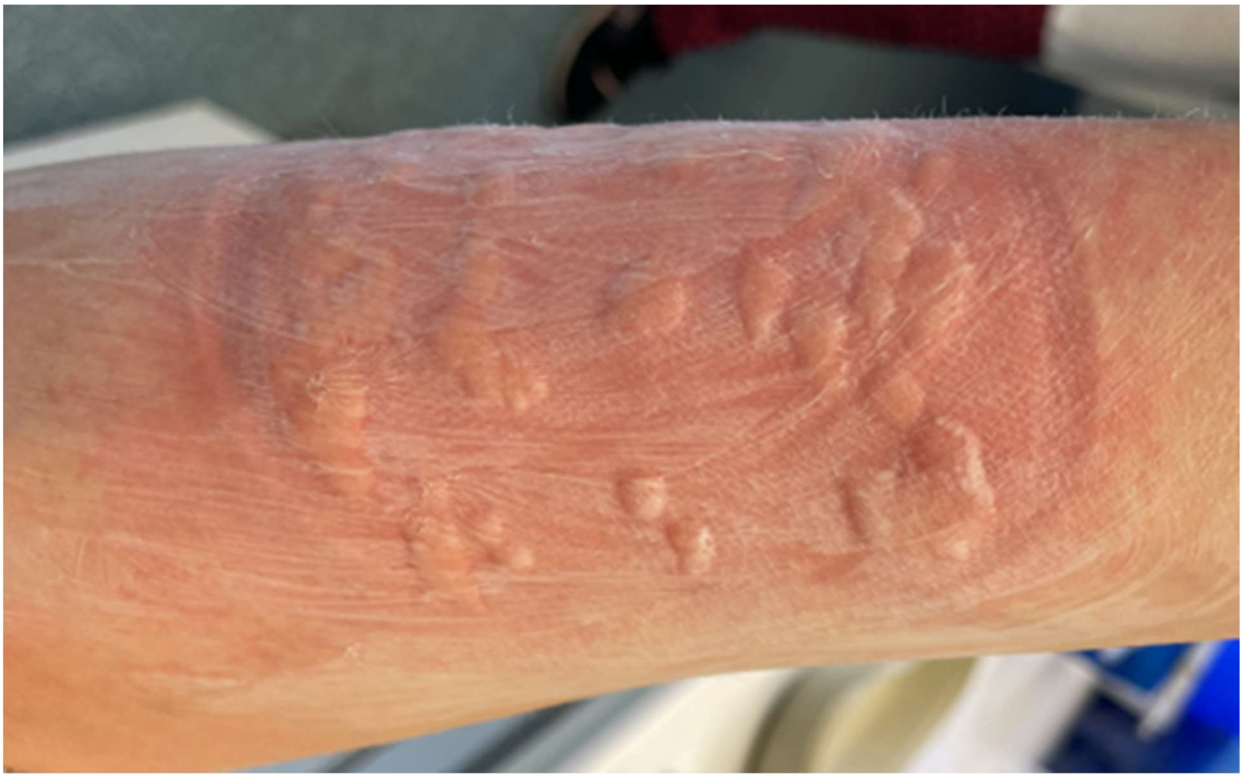

**Table S2.** Overview of all unsolicited AEs with their preferred MedDRA terms.
AE duration is expressed in decimal hours, calculated as the total number of minutes between AE onset and end, divided by 60 (e.g. a duration of 360 hours and 13 minutes is displayed as 360,22).
*Grading was based on The Division of AIDS (DAIDS) Table for Grading the Severity of Adult and Pediatric Adverse Events, Corrected Version 2.1 (July 2017)
Δ There was no grade available in the AIDS criteria for these AEs, grading supplied is based on clinical assessment

| Participant Id | Preferred MedDRA terms | Duration (in hours) | Grading | Causality |
|---|---|---|---|---|
| 3C | 10020407 (Hot flashes) | 68 | moderate | Probably related |
| 1D | 10028411 (Myalgia) | 121 | mild | Not related |
| 3B | 10041367 (Sore throat) | 41 | mild | Not related |
| 2D | 10046306 (Upper respiratory tract infection) | 135,5 | mild | Not related |
| 1D | 10024891 (Low back pain) | 53,5 | mild | Not related |
| 3D | 10016373 (Feelings of weakness) | 17 | mild | Definitely related |
| 3C | 10077974 (Peripheral neuropathic pain) | 6 | moderate | Unlikely related |
| 1C | 10046306 (Upper respiratory tract infection) | Still present at end of follow-up | mild | Not related |
| 1D | 10041367 (Sore throat) | 42,5 | moderate | Unlikely related |
| 1A | 10046306 (Upper respiratory tract infection) | 87 | mild | Not related |
| 3C | 10019211 (Headache) | Still present at end of follow-up | mild | Unlikely related |
| 2C | 10039296 (Runny nose) | 43,83 | mild | Not related |
| 2B | 10017924 (Gastroesophageal reflux) | 64 | moderate | Probably related |
| 1C | 10039296 (Runny nose) | Still present at end of follow-up | mild | Not related |
| 2D | 10087685 (Menstrual headache) | 9 | mild | Not related |
| 1A | 10045148 (Tummy ache) | 83 | mild | Probably related |
| 1D | 10028411 (Myalgia) | 37,75 | moderate | Possibly related |
| 1B | 10045148 (Tummy ache) | 189 | mild | Probably related |

| | | | | |
|---|---|---|---|---|
| 1C | 10028411 (Myalgia) | 12,25 | mild | Unlikely related |
| 3C | 10011224 (Cough) | 48 | mild | Unlikely related |
| 3A | 10000659 (Acute allergic urticaria) | 23,75 | severe | Definitely related |
| 2B | 10023082 (Itch) | 264 | moderate | Definitely related |
| 1A | 10024891 (Low back pain) | 408 | mild | Unlikely related |
| 2C | 10046306 (Upper respiratory tract infection) | 48 | mild | Not related |
| 1C | 10046306 (Upper respiratory tract infection) | 42 | mild | Not related |
| 3C | 10038986 (Retrosternal chest pain) | 2 | moderate | Unlikely related |
| 3D | 10045148 (Tummy ache) | 73 | moderate | Probably related |
| 1D | 10041367 (Sore throat) | 288 | mild | Unlikely related |
| 2B | 10046306 (Upper respiratory tract infection) | 25 | mild | Not related |
| 1D | 10087685 (Menstrual headache) | 63,5 | moderate | Not related |
| 1B | 10028411 (Myalgia) | 113 | mild | Not related |
| **Participant Id** | **Laboratory AEs** | **Duration (in hours)** | **Grading*** | **Causality** |
| 3C | 10011418 (CRP increased) | 360,22 | Δ mild | Probably related |
| 1C | 10003544 (AST increased) | 266,05 | mild | Probably related |
| 1C | 10001551 ( ALT increased) | 266,05 | mild | Probably related |
| 1A | 10065393 (Absolute neutrophil count increased) | 258,17 | Δ mild | Possibly related |
| 3D | 10001551 (ALT increased) | 505 | mild | Probably related |
| 1B | 10024384 (Leukopenia) | 411,72 | mild | Probably related |
| 2B | 10024384 (Leukopenia) | 406,42 | mild | Probably related |
| 1A | 10024384 (Leukopenia) | 335,85 | mild | Probably related |
| 2C | 10011418 (CRP increased) | 382,05 | mild | Probably related |
| 3B | 10005851 (Blood urea increased) | 336.48 | mild | Unlikely related |
| 3D | 10017670 (Gamma GT increased) | 309,25 | Δ mild | Probably related |

| | | | | |
|---|---|---|---|---|
| 2A | 10011418 (CRP increased) | 382,07 | Δ mild | Definetly related |
| 1D | 10003544 (AST increased) | 333,15 | mild | Possibly related |
| 3D | 10011418 (CRP increased) | 409,15 | Δ mild | Probably related |
| 3D | 10024051 (LDH increased) | 409,15 | Δ mild | Probably related |
| 3C | 10024384 (Leukopenia) | 360,22 | mild | Probably related |
| 2A | 10017670 (Gamma GT increased) | 792,57 | Δ mild | Probably related |
| 3C | 10024051 (LDH increased) | 360,22 | Δ mild | Probably related |
| 3D | 10001551 (ALT increased) | 167,77 | mild | Probably related |
| 2C | 10024051 (LDH increased) | 382,05 | Δ mild | Probably related |
| 3B | 10011418 (CRP increased) | 359,75 | Δ mild | Probably related |
| 1B | 10003544 (AST increased) | 266,3 | moderate | Probably related |
| 1B | 10001551 (ALT increased) | 266,3 | moderate | Probably related |
| 3A | 10001551 (ALT increased) | 336,35 | mild | Not related |
| 1D | 10024384 (Leukopenia) | 314,47 | mild | Probably related |
| 3A | 10004685 (Bilirubin conjugated increased) | 118,78 | mild | Possibly related |
| 3A | 10056806 (Bilirubin total increased) | 118,78 | mild | Possibly related |
| 3A | 10003544 (AST increased) | 336.35 | moderate | Unlikely related |
| 3D | 10001551 (ALT increased) | 72,15 | moderate | Probably related |
| 2A | 10003544 (AST increased) | 1174,63 | mild | Definetly related |
| 2A | 10001551 (ALT increased) | 1174,63 | mild | Definetly related |
| 3C | 10004685 (Bilirubin conjugated increased) | 70,73 | mild | Probably related |
| 3C | 10056806 (Bilirubin total increased) | 70,73 | mild | Probably related |
| 1A | 10048553 (Leukocyte count increased) | 258,17 | Δ mild | Possibly related |
| 3D | 10003544 (AST increased) | 744,92 | mild | Probably related |
| 3C | 10003544 (AST increased) | 698,55 | mild | Probably related |

| | | | | |
|---|---|---|---|---|
| 3C | 10001551 ( ALT increased) | 698,55 | mild | Probably related |
| 3D | 10024384 (Leukopenia) | 409,15 | mild | Probably related |
| 2B | 10003544 (AST increased) | 406.42 | mild | Probably related |
| 2B | 10011418 (CRP increased) | 741,95 | Δ mild | Probably related |
| 2B | 10024051 (LDH increased) | 406,42 | Δ mild | Probably related |
| 3A | 10011418 (CRP increased) | 360,2 | Δ mild | Probably related |
| 3A | 10024051 (LDH increased) | 336,35 | Δ mild | Unlikely related |
| **Participant Id** | **Vital signs AEs** | **duration (in hours)** | **Grading*** | **Causality** |
| 1C | 10014474 (Elevated blood pressure reading without diagnosis of hypertension) | 23,95 | mild | Not related |
| 2B | 10012753 (Diastolic blood pressure increased) | 37,5 | mild | Probably related |
| 3C | 10012753 (Diastolic blood pressure increased) | 23,42 | mild | Unlikely related |
| 3C | 10014474 (Elevated blood pressure reading without diagnosis of hypertension) | 122,5 | mild | Not related |
| 2B | 10012753 (Diastolic blood pressure increased) | 23 | mild | Probably related |
| 1D | 10021113 (Hypothermia) | 25,13 | Δ mild | Not related |
| 3A | 10021113 (Hypothermia) | 24 | Δ mild | Not related |
| 3C | 10012753 (Diastolic blood pressure increased) | 166,08 | mild | Unlikely related |
| 2A | 10043071 (Tachycardia) | 456,5 | Δ mild | Not related |
| 2B | 10021113 (Hypothermia) | 22,5 | Δ mild | Not related |
| 1D | 10043071 (Tachycardia) | Still present at end of follow-up | Δ mild | Not related |

**Figure S4.** Line plot showing the median cohort trajectories of AST and ALT counts over five key study timepoints.

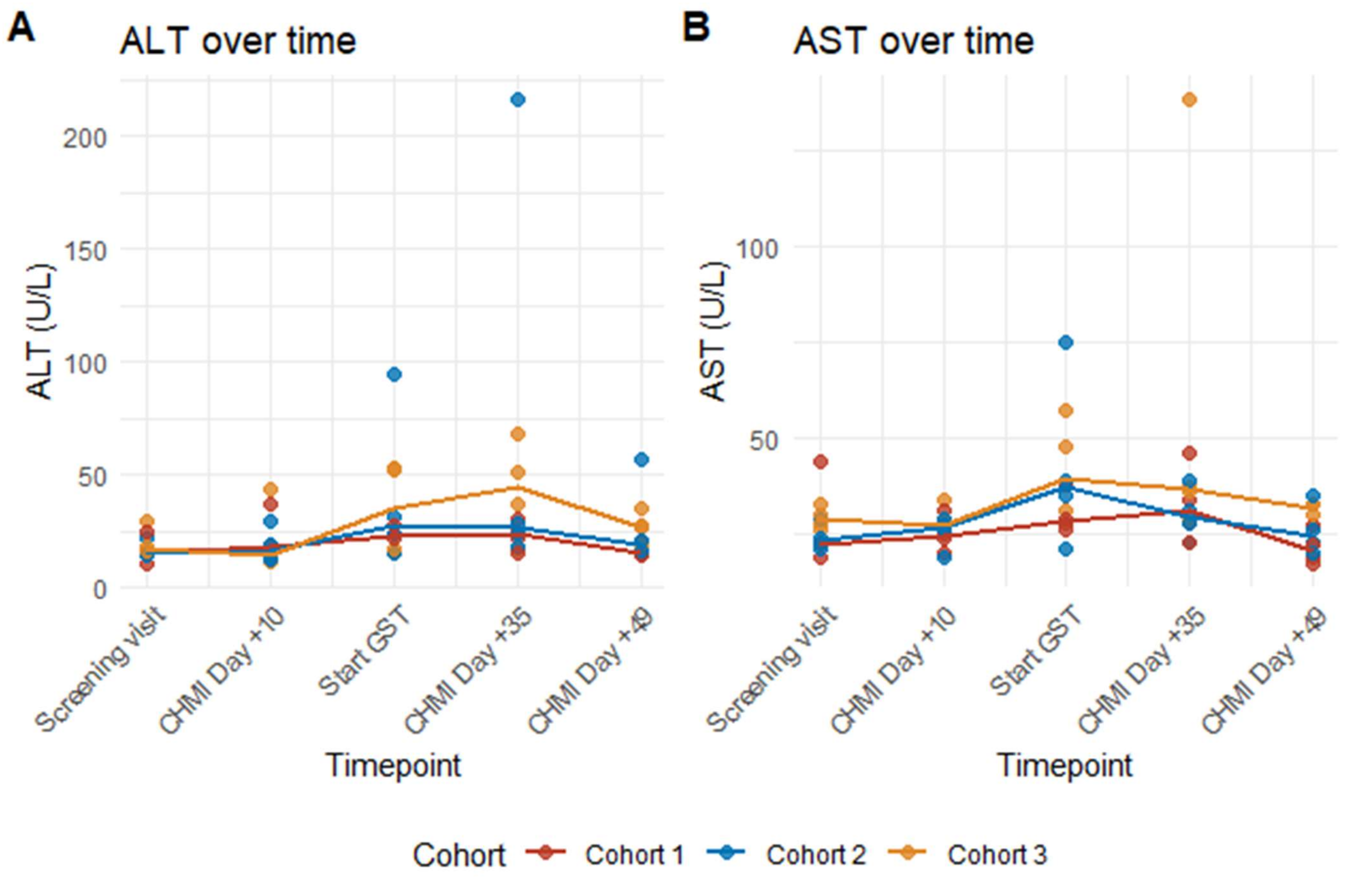

### Thick blood smear preparation, staining and quantification SOP

#### TBS preparation

1. Create a paper template for the circumference of the thick blood smear, this should be the diameter of a one euro cent coin. The template can be prepared using an inkpad and the coin as a stamp (can be re-used).
2. Put on a lab coat and gloves.
3. Turn on the heat block to 37 °C.
4. Label two glass slides with the participant's identification number, visit day (i.e. CHMI+5) and date.
5. Place the glass slide on the paper template.
6. If using blood from EDTA tube, gently invert the tube being careful not to create bubbles.
7. Take just a bit more than 15 µl of blood using a calibrated pipette. Check that there are no bubbles in the tip of the pipette.
8. Pipette the blood in the middle of the circle by pressing the pipette until **the first stop**. Making sure not to press it down completely, there should still be some blood left in the tip!
   Keep the tip of the pipette on the glass slide and spread the blood to fill the circle by making circular motions outwards.
9. Once the circle is full return the tip of the pipette to the middle without taking it from the slide. Once in the middle remove it from the glass slide without pressing it down completely.
10. Repeat step 7-9 till you have four thick blood smears on two glass slides (the amount can differ per protocol).
11. Once a glass slide is done put it on the heat block for a minimum of 15 minutes (longer is better).

#### Preparation of Giemsa solution

Note: <u>Always create the Giemsa solution fresh on the day of use. Do not use if older than 6 hours.</u>

1. Put on a lab coat and gloves.
2. Create a safe working space, the Giemsa stain should always be made in a well-ventilated area (for example under an extractor hood) in a space that can be cleaned easily if the stain spills (for example put the coloring container in a glass beaker).
3. Gently invert the Giemsa AEM stock solution
4. Using a disposable 1 ml pipette (in a pipetteboy), pipette 0.8 ml of Giemsa AEM stock into the coloring container.
5. Using a disposable 10 ml pipette, pipette 10 ml of **cold** tap water into the coloring container (make sure the water is cold! Lukewarm or hot water will change the color of the staining). Keep the pipette.
6. Using the same 10 ml pipette, pipette 10 ml of Giemsa buffer into the coloring container and resuspend the solution well.

#### Staining

<u>Note: Keeping temperature during drying of the blood below 45°C is critical. Never heat over 45°C!</u>

1. Before putting the glass slide in the Giemsa stain make sure that it has been on a heat block of 37 °C for a minimum of 15 minutes and that the thick blood smear is dry.
2. Put on a lab coat and gloves.

3. Put the glass slide in the freshly prepared Giemsa stain for **30 minutes** (the time may be adapted to ensure optimal staining).
4. Prepare two containers with cold tap water (make sure the water is cold! Lukewarm or hot water will change the color of the staining). Make sure that the containers are big enough to horizontally fit the glass slide.
5. Carefully remove the glass slide from the Giemsa stain using a tweezer.
6. Put the slide horizontally in the first water container and gently move it from side to side for about 5 seconds.
7. Remove it from the first water container and put the glass slide in the second container, repeating step 6.
8. Remove any excess water still clinging to the **bottom** of the slide with a paper towel. If the top of the glass slide is still very wet you can drain the excess water by placing the object glass upright on a tissue for a few moments. Make sure not to touch the thick blood smears!
9. Put the glass slide on the heat block (37 °C) for a minimum of 7 minutes.
10. Check if the thick blood smear is dry. If yes, the thick blood smear is ready for reading.

**Microscopy**

- Choose an area of the TBS where blood is evenly distributed to examine with a 100x oil immersion objective.
- Apply a small drop of immersion oil to an area of the TBS that appears mauve colored (here a good color balance between the blue and red stains is present).
- The minimal number of parasites to be seen to declare a slide positive is 2.
- Examine at least 10 microscope fields, if the parasite count is > 50 parasites per field.
- Examine at least 20 microscope fields, if the parasite count is between 2 and 50 parasites per field.
- If, on average, less than 1 parasite is present per field read until at least 10 parasites are counted or 215 fields without parasite have been observed (this corresponds to a 95% probability to detect at least 10 parasites per microliter with the condition to detect at least 2 parasites).
- In case one parasite has been observed in 215 fields, reading up to a total of 430 fields shall be done.

All TBS must be read by at least two independent microscopists including at least one microscopist who passed the slide reading qualification (Qualified Microscopists (QMi)). A third QMi reading is required when results among first two readers are discordant.

**Calculation of parasite density**

The number (No) of parasites per microliter blood is calculated as:

$$\frac{No\ of\ parasites}{No\ of\ fields} \times MF = parasites\ per\ \mu l.$$

E.g. 24 parasites in 20 fields and a MF of 454 equals 545 parasites/µl; 2 parasites in 200 fields at the same MF are equivalent to 5 parasites/µl.
